# Scalable Deep Learning of Histology Images Reveals Genetic and Phenotypic Determinants of Adipocyte Hypertrophy

**DOI:** 10.1101/2025.02.11.25322053

**Authors:** Emil Jørsboe, Phil Kubitz, Julius Honecker, Andrea Flaccus, Dagmar Mvondo, Matthias Raggi, Craig A. Glastonbury, Torben Hansen, Matthias Blüher, Aleksander Krag, Hans Hauner, Philip D. Charles, Cecilia M. Lindgren, Christoffer Nellåker, Melina Claussnitzer

**Affiliations:** Big Data Institute, Li Ka Shing Centre for Health Information and Discovery, University of Oxford, Oxford, United Kingdom; Nuffield Department of Population Health, University of Oxford, Oxford, United Kingdom; Novo Nordisk Foundation Center for Basic Metabolic Research, Faculty of Health and Medical Sciences, University of Copenhagen, Copenhagen, Denmark; Center for Liver Research, Department of Gastroenterology and Hepatology, Odense University Hospital, Odense, Denmark; Technical University of Munich, Else Kröner-Fresenius-Center for Nutritional Medicine, Chair of Nutritional Medicine, School of Life Sciences, Gregor-Mendel-Straße 2, 85354, Freising-Weihenstephan, Germany; Broad Institute of MIT and Harvard, Medical and Population Genetics Program & Type 2 Diabetes Systems Genomics Initiative, Cambridge, MA, USA; Novo Nordisk Foundation Center for Genomic Mechanisms of Disease, Broad Institute of MIT and Harvard, Cambridge, MA, USA; Institute of Nutritional Sciences, University of Hohenheim, Stuttgart, Germany; Department of General and Visceral Surgery, Karl-Olga-Krankenhaus, Stuttgart, Germany; Nuffield Department of Medicine, University of Oxford; Oxford, United Kingdom; Human Technopole, Viale Rita Levi-Montalcini 1, 20157, Milan, Italy; Helmholtz Institute for Metabolic, Obesity and Vascular Research (HI-MAG) of the Helmholtz Zentrum München at the University of Leipzig and University Hospital Leipzig, Leipzig, Germany; Medical Department III - Endocrinology, Nephrology, Rheumatology, University of Leipzig Medical Center, Leipzig, Germany; Institute of Clinical Research, University of Southern Denmark, Odense, Denmark; Institute for Nutritional Medicine, School of Medicine and Health, Technical University of Munich, Georg-Brauchle-Ring 62, 80992, Munich, Germany; Target Discovery Institute, Centre for Medicines Discovery, Nuffield Department of Medicine, University of Oxford, Oxford, United Kingdom; Nuffield Department of Women’s & Reproductive Health, University of Oxford, Oxford, United Kingdom; Diabetes Unit and Center for Genomic Medicine, Massachusetts General Hospital, Boston, MA, USA

## Abstract

**Background:** White adipose tissue dysfunction has emerged as a critical factor in cardiometabolic disease development, yet the cellular microstructure and genetic architecture of adipocyte morphology remain poorly explored.

**Methods:** We introduce Adipocyte U-Net 2.0, an advanced deep learning method for the semantic segmentation of adipose tissue histology, enabling analysis of over 27 million adipocytes from 2,667 individuals.

**Findings:** Our approach revealed that adipocyte hypertrophy associates with metabolic dysfunction, including increased fasting glucose, glycated hemoglobin, leptin, and triglycerides, with decreased adiponectin and HDL cholesterol levels. Through the largest genome-wide association study of adipocyte size to date (N_Subcutaneous_ = 2,066, N_Visceral_ = 1,878), we identified four genome-wide significant loci: two in sex-combined analysis (rs73184721 in *NAALADL2* and rs200047724 in *NRXN3*) and two female-specific variants (rs140503338 and rs11656704 in *ULK2*). Notably, these genetic associations showed congruent relationships with cardiometabolic traits, suggesting shared biological mechanisms.

**Interpretation:** Our findings demonstrate the utility of deep learning for adipocyte phenotyping at scale and provide novel insights into the genetic basis of adipocyte morphology and its relationship to metabolic disease.

## Introduction

Obesity is a rapidly growing global healthcare problem. Individuals with obesity are characterised by an increased cardiometabolic disease risk, including type 2 diabetes (T2D), coronary artery disease, and metabolic dysfunction–associated steatotic liver disease (1,2). Recent large-scale genome-wide association studies (GWAS) of body fat distribution, fasting insulin levels, and T2D highlight white adipose tissue (WAT) as a key tissue in which disease-associated variants manifest their effect (3).

During the development of obesity, WAT expands due to a combination of increasing adipocyte size (hypertrophy) and number of adipocytes (hyperplasia). While WAT expansion through hyperplasia is not associated with obesity-associated metabolic complications, hypertrophy is associated with ectopic fat accumulation in the liver, skeletal muscle, pancreas, and heart tissue, local inflammation (4), cardiometabolic risk (4), and impaired glucose metabolism (5). Expansion of white adipose tissue in adults is primarily characterised by hypertrophy (6). In individuals with obesity and adipocyte hypertrophy, the storage capacity of triglycerides is limited, and further caloric overload leads to fat accumulation in peripheral tissues and in visceral adipose depots (4). Enlargement of adipocytes is associated with low-grade chronic inflammation, insufficient angiogenesis, and excessive collagen deposition, which further leads to dysfunctional adipokine release (7). Adipocyte hypertrophy correlates strongly with insulin resistance and T2D in visceral adipose tissue, independently of body mass index (BMI) (8).

Obesity and fat distribution are highly heritable traits (9,10), and recent large-scale genetic association studies (11,12) have identified well over 1,000 independent genetic risk loci for fat mass and distribution-related traits. However, the disease-driving cellular processes remain largely unclear. We hypothesized that adipocyte hypertrophy is one such disease-driving phenotype. There is a lack of large-scale, well-powered GWAS mapping of the genetic determinants of adipocyte morphology (13,14), and the underlying genes and regulatory pathways involved in the size of adipocytes are not well characterised (15).

Historically, a practical limiting factor has been the lack of tools that allow for scalable and affordable mapping of adipocyte size. WAT histology samples can be used for studying adipocyte morphology (16), but manual annotation is cumbersome and extremely slow, and software for image-based measurements of adipocyte rarely scales well for practical application to large datasets. We previously developed Adipocyte U-Net, a deep learning approach for estimating the size of adipocytes across thousands of white adipose tissue histology samples (17). This model was validated against established methods, including Adiposoft (16) and CellProfiler (17), but analysis of contemporary data cohorts (N ∼ 1,000) did not identify genome-wide significant genetic associations, likely due to limited statistical power, cohort heterogeneity, and limits on phenotypic resolution.

A limitation of our previous approach was the image-tiling strategy, which involved splitting whole slide images (WSIs) into sub-images and analyzing only tiles containing complete adipocytes. This methodology excluded adipocytes intersecting tile boundaries, which may bias size estimates downward since larger hypertrophic cells are more likely to intersect a boundary. Our previous U-net-based size estimates were consistently smaller compared to those from Adiposoft or CellProfiler (14).

To address this limitation and enable a comprehensive investigation of adipocyte morphology and its genetic determinants, we have developed an improved deep learning approach, Adipocyte U-Net 2.0, capable of accurately segmenting adipocytes across entire WSIs while properly accounting for cells that cross tile boundaries. Applying this enhanced methodology to an unprecedented collection of 9,000 WSIs containing over 27 million adipocytes from 2,667 individuals across five independent cohorts, we characterize the relationship between adipocyte hypertrophy and metabolic health, and perform the largest GWAS of adipocyte size to date, revealing novel genetic determinants of this clinically relevant cellular phenotype.

## Methods

### Training Adipocyte U-Net 2.0

We implemented Adipocyte U-Net 2.0 based on U-net (18) in PyTorch (19) for the semantic segmentation of adipocytes in WSIs. The model is based on the design in (17), with added dilations for the lowest resolution layer. The training, validation and hold-out test split was 70%, 15% and 15%, respectively.

The dice score of our validation with the lowest loss (0·36) was 0·83 (Supplementary Figure 1), the dice score of our held-out test set was 0·87.

### Deep learning-derived phenotypes

For prediction, we adapted the code base for HAPPY (20), implemented in Python and using PyTorch for semantic segmentation. Tiles of 1024×1024 pixels are used, starting from the top left corner of the WSI and moving left to right in rows (Figure 1).

**Figure 1.**
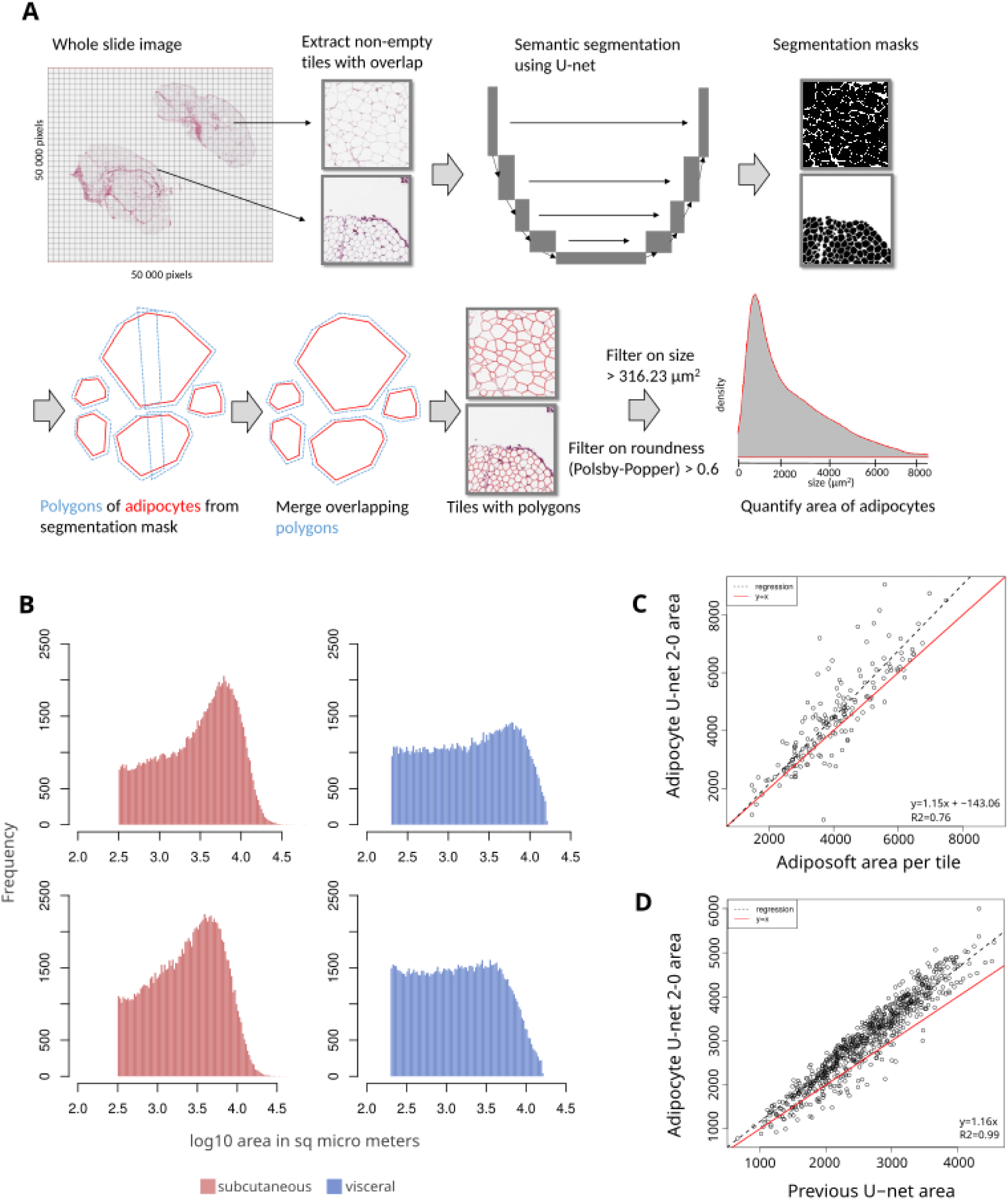
Adipocyte U-Net 2.0 captures adipocyte size-derived phenotypes from WSIs at scale. **A)** Workflow of adipocyte size quantification using Adipocyte U-net 2.0. **B)** 500 randomly sampled segmented adipocytes from WSIs from the Munich cohort using the filters from Adipocyte U-net 2.0 (upper histograms) using the filters (lower histograms) from (17). **C)** Comparison of mean adipocyte size estimates from Adipocyte U-net 2.0 and Adiposoft (28) based on 1024×1024 pixel tiles from the Leipzig cohort. **D)** Comparison of mean adipocyte size estimates from Adipocyte U-net 2.0 and (17) based on WSIs from the GTEX cohort.

Segmentation is performed twice on each WSI, with a pixel size of 0·2500 μm and 0·5034 μm. The segmentation mask is efficiently stored as polygons, using the shapely library.

### Post-Processing

The merged polygon list was filtered for area greater than 316·23 μm^2^ and a Polsby-Popper (PP) roundness value (21) greater than 0·6 (empirically based distribution derived thresholds; prefiltered distributions and selected thresholds are shown in Supplementary Figure 2 & 3) (Figure 1A).

From the filtered polygons from each individual, we calculated mean adipocyte size and upper 95%-quantile of adipocyte size, and the ratio between mean adipocyte size in visceral and subcutaneous visceral adipose tissue.

For more technical details on the training of the model, derivation of phenotypes and post-processing see the Supplementary Methods.

### Validation of Adipocyte U-Net 2.0-based estimates of adipocyte size against Adiposoft

Using WSIs from the Leipzig cohort, we ran Adiposoft on 10 tiles (1024×1024 pixels) per 17 WSIs. The WSIs were chosen randomly, and the tiles containing adipocytes were chosen. We used the same lower size cutoff of 316·23 μm^2^ as for our Adipocyte U-net 2.0-based approach and adipocytes at the edge of the tile were discarded, as Adiposoft only includes the area inside the tile for adipocytes at the edge.

For comparison with our Adipocyte U-net 2.0, we extracted the polygons within the coordinates of the tiles used with Adiposoft, we also removed polygons touching the edge of the tile to ensure consistent comparison.

### Cohorts

WAT samples from subcutaneous and visceral depots were obtained from five different study cohorts (Table 1). Each study participant gave written informed consent and the protocols were approved by the corresponding ethics committee. Munich and Hohenheim: 5716/13, 1946/07, 409/16s. Leipzig: 159-12-21052012, 017-12-23012012. ENDOX: REC: 09/H0604/58, IRAS: 8282, GTEX: NIH (project id phs000424.v7.p2).

**Table 1.** An overview of the five study cohorts used in this study and the number of WSIs per cohort. BMI, age and sex information is the number of individuals with phenotypes; these numbers might therefore differ from the number of individuals with WSIs.

| Cohort | Individuals with WSI (SC / VC) | Phenotypes | BMI (mean, SD) | Age (mean, SD) | Sex (M / F) |
| --- | --- | --- | --- | --- | --- |
| Munich | 211 / 232 | Anthropometric and metabolic | 45·13, 12·58 | 46·13, 12·71 | 105 / 204 |
| Leipzig | 1151 / 1145 | Anthropometric and metabolic | 46·62, 11·49 | 49·77, 13·01 | 403 / 766 |
| Hohenheim | 47 / 50 | Anthropometric and metabolic | 46·55, 6·23 | 42·31, 13·09 | 37 / 114 |
| Endox (Oxford) | 425 / 55 | Anthropometric | 26·60, 5·80 | 32·78, 7·59 | 0 / 352 |
| GTEX | 918 / 759 | Anthropometric and metabolic | 27·32, 4·13 | 52·76, 12·91 | 653 / 327 |

### Munich, Leipzig and Hohenheim cohorts

Male and female patients undergoing abdominal laparoscopic surgery were included. Subcutaneous adipose tissue biopsy derived from beneath the skin at the abdominal surgical incision site and visceral adipose tissue was obtained at the proximity of the angle of His. After sampling, approx. 5 mm^3^ pieces of the tissue biopsies were sectioned and fixed in 4% paraformaldehyde for histology.

### ENDOX cohort

Female premenopausal participants were recruited at the Oxford Endometriosis CaRe Centre between 2012 and 2018 for endometriosis screening via laparoscopy. Biopsies of the endometrium, peritoneal fluid, and adipose tissue from subcutaneous and visceral depots were obtained (17).

### GTEX cohort

Whole slide images from HE stained subcutaneous and visceral adipose tissue samples were obtained from the Genotype Tissue and Expression (GTEX) project. Samples were obtained from individuals post-mortem. Subcutaneous adipose tissue biopsies were sampled at the leg, approx. 2 cm below the patella, and visceral adipose tissue was obtained from the omentum (22).

### Histology and imaging

Dehydration and clearing of fixated histology samples were performed automatically (TP1020, Leica, Germany). Afterwards, samples were embedded in paraffin and 5 μm thick tissue sections were obtained using a rotary microtome (RM2255, Leica, Germany). Subsequently, the tissue sections were transferred to a glass microscope slide, hematoxylin and eosin (H&E) staining was applied using a fully automated multistainer (ST5020, Leica, Germany) and the samples were immediately coverslipped. Digital whole slide images were obtained using a slide scanner (Aperio AT2, Leica, Germany).

### Genotyping

The Leipzig cohort was genotyped in two different batches. Lack of batch effect was confirmed via PCA with pruning and only with autosomal markers, filtering for MAF>0·05 and missingness<0·01 (Supplementary Figure 4). The two genotype files were merged into one file, which was used for all subsequent analyses.

For an overview of the genetic data and quality control, see respectively Supplementary Table 1 and Supplementary Methods.

### Imputation

Each SNP-chip was phased and imputed independently after applying a quality control filter (see “Genetic Quality Control” section in Supplementary Methods), splitting by chromosome, and preparing the input files according to the TOPMed guidelines (https://topmedimpute.readthedocs.io/en/latest/). The TOPMed Imputation server was used (23). Eagle v2.4 (24) was used for phasing and Minimac4 (23) for imputation, both with the multiethnic TOPMed r2 reference panel, selecting the “vs. TOPMed panel” as the population.

### Association analysis

GWAS was performed using bimbam mean genotype files, including genetic variants with an imputation Rsq > 0·3. A linear mixed model was run on each cohort using GEMMA (25). The model was adjusted for sex, age, age^2^, sex⋅age, and the top ten genetic PCs. The ENDOX cohort was analysed jointly and the linear mixed model was additionally adjusted for which SNP-chip type. The Hohenheim cohort and part of the Munich cohort genotyped on the same SNP-chip were jointly analysed. The phenotypes were analysed raw and inverse normal quantile transformed. The outputs from each cohort were meta-analysed and the joint estimates of effect size and P-value were derived using the inverse variance-based strategy using METAL (26).

Furthermore, a phenome-wide association study (PheWAS) was performed for the lead variants using a meta-analysis-based approach. We focused on traits that were linked to hypertrophy, i.e. BMI, waist-hip ratio (WHR), plasma glycated hemoglobin, fasting plasma glucose, fasting plasma insulin, plasma 2-hour blood glucose, Interleukin 6, Adiponectin, type II diabetes status and C-reactive protein.

### Colocalisation

Colocalisation between adipocyte-derived and other metabolic traits was assessed using a Bayesian statistical test with the coloc R-package (version 5.2.2) (27). Genomic regions were defined by windows of 1 Mb around suggestive and significant GWAS signals (P < 10^−5^). Overlapping regions were merged. The minimal data required by coloc were used as input: SNP IDs, positions, effect sizes, standard errors, data type (quantitative or case-control), MAF and sample sizes. To identify potential causal variants, 95% credible sets of variants based on their posterior inclusion probabilities were created. A posterior probability PPH4 (H4: associations with both traits due to a single causal variant) > 80% was considered sufficient for the identification of colocalisation.

We analysed all the traits listed in Figure 2.

**Figure 2.**
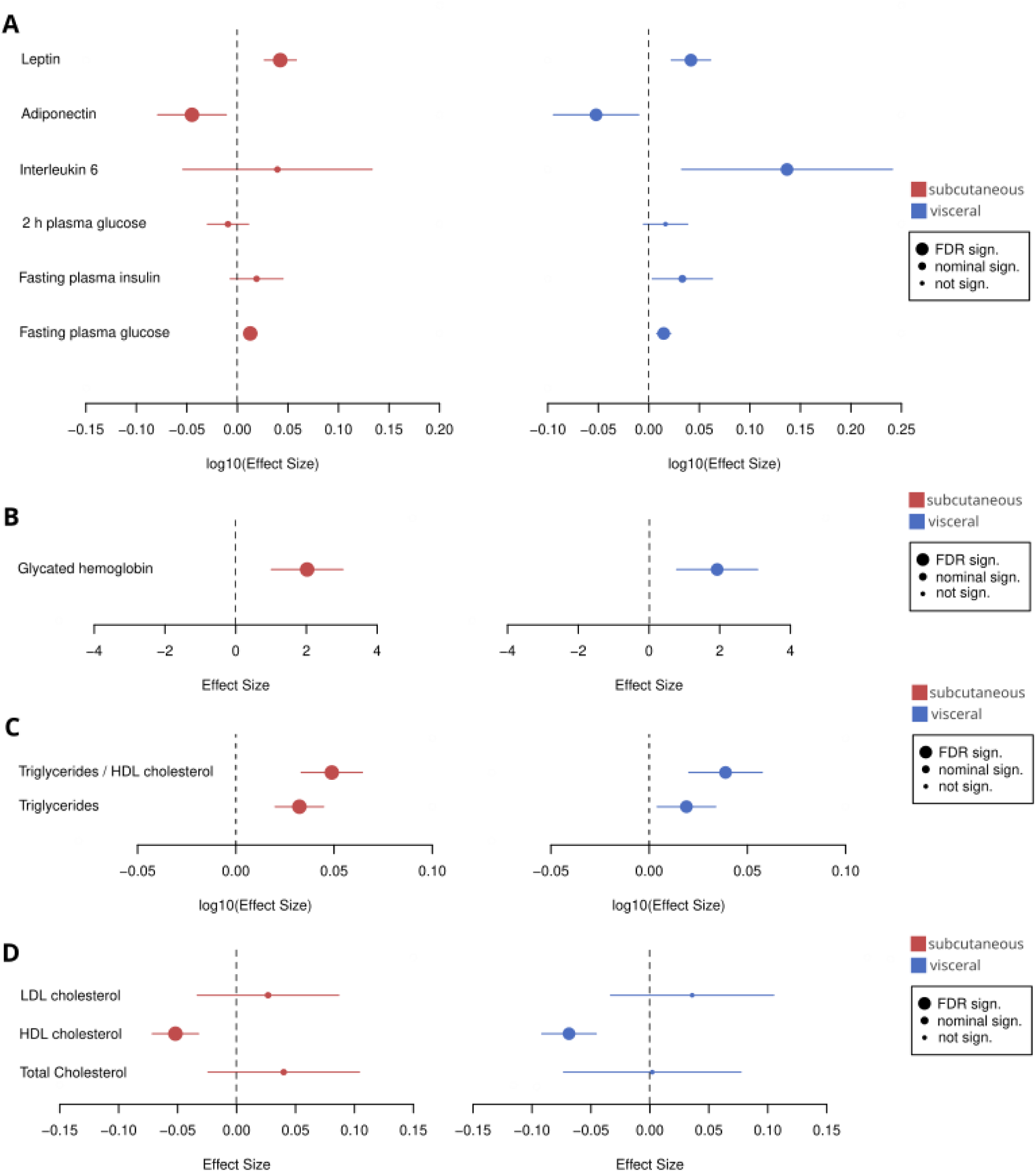
Effect size of mean adipocyte size on adipokine and glycemic traits **A)** and **B)** and lipid traits **C)** and **D)** with the 95% confidence intervals. q-values for the False Discovery Rate (FDR) are 0·036 for **A)** and **B)** and 0·03 for **C)** and **D)** respectively.

### Adipocyte hypertrophy in the Leipzig cohort

Phenotype associations with mean adipocyte size were tested using a linear model. Adipocyte size was divided by 1,000 to make the effect sizes more meaningful. The linear model was adjusted for age, sex, BMI and T2D:

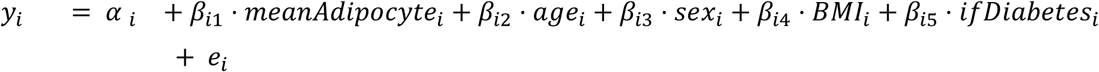

Details on estimating the effect of subcutaneous adipose tissue sampling location and adipocyte size epidemiology using linear mixed models are described in the Supplementary Methods.

## Results

To facilitate large-scale analysis of adipocyte morphology, we developed Adipocyte U-Net 2.0, an advanced deep learning framework for the semantic segmentation of adipocytes in histological whole slide images (WSIs). Our improved methodology builds upon our previous work to implement a comprehensive approach to WSI analysis that eliminates potential size bias. Rather than processing discrete, non-overlapping image tiles independently, Adipocyte U-Net 2.0 performs semantic segmentation across entire WSIs using overlapping tiles, enabling accurate identification and measurement of all adipocytes, including those that cross tile boundaries (Figure 1A).

From the segmented adipocytes, we calculated five key phenotypes for subsequent analyses: mean adipocyte size, upper 95%-quantile of adipocyte size (as a proxy for hypertrophy), and the ratio between mean adipocyte size in visceral and subcutaneous adipose tissue. Through optimized parallelization, our method achieved unprecedented throughput with a runtime of one week, enabling the analysis of 9,000 WSIs containing over 27 million adipocytes from 2,667 individuals across five independent cohorts.

### The Adipocyte U-net 2.0 improves the detection of hypertrophic adipocytes compared to previous models

We validated Adipocyte U-net 2.0 against both our previous model and the semi-automatic segmentation via Adiposoft (28). Adipocyte U-net 2.0-based estimates of mean adipocyte size were benchmarked against Adiposoft, showing strong agreement between methods with a linear correlation of R^2^ = 0·76 (Figure 1C), confirming the accuracy of our deep learning approach. Comparing Adipocyte U-net 2.0 against our previous model, using identical filtering criteria, we observe a strong linear correlation comparing the adipocyte size estimates from both models (R2_subcutaneous_ = 0·99, R^2^_visceral_ = 0·99) (Figure 1D). Importantly, however, the current implementation of the Adipocyte U-net 2.0 results in consistently larger size estimates (β_subcutaneous_ = 1·44, β_visceral_ = 1·4). Additionally, the model enables the detection of very large adipocytes at the upper tail-end of the distribution (Figure 1B). Our results demonstrate that Adipocyte U-Net 2.0 maintains the speed and accuracy advantages of our previous approach while addressing limitations in the detection of very large adipocytes.

### Adipocyte hypertrophy is associated with demographic and metabolic status predominantly in visceral adipocytes

We set out to investigate the relationship between adipocyte size and a series of clinical variables across adipose tissue samples from 2215 visceral and 2527 subcutaneous donors with WSIs, encompassing 27 million adipocytes.

Mean adipocyte size increased with BMI in both the visceral depot (μ = 0·39, P_μ_ = 1·89⋅10^−7^, *τ^2^*= 0·021, P*_τ_* = < 0·0001, *I^2^*= 88·5%) and the subcutaneous adipose depot (μ = 0·40, P = 8·28⋅10^−21^, *τ^2^*= 0·0057, P*_τ_* = 0·0053, *I^2^*= 72·8%) (Supplementary Figure 5-6). Mean adipocyte size also increased with age in the visceral adipose depot (μ = 0·12, P_μ_ = 0·00014, *τ^2^* = 0·0020, P*_τ_* = 0·09, *I^2^*= 51%), but not in subcutaneous (μ = 0·07, P = 0·15, *τ^2^* = 0·0068, P*_τ_* = 0·0005, *I^2^* = 79·9%) (Supplementary Figure 7-8). Stratifying by sex, mean adipocyte size was smaller in females compared to males in visceral adipose tissue (μ = −0·32, P_μ_ = 6·77⋅10^−15^, *τ^2^*= 0, P*_τ_* = 0·49), this sex-specificity of adipocyte size was not detected in the subcutaneous depots (μ = 0·12, P_μ_ = 0·41, *τ^2^*= 0·061, P*_τ_* = < 0·0001, *I^2^* = 91·3%) (Supplementary Figure 9-10).

Comparing T2D cases to controls, we observed an increase in mean adipocyte size in the visceral depot (μ = 0·11, P_μ_ = 0·021, *τ^2^* = 0, P*_τ_* = 0·69) (Supplementary Figure 11-12). Mean visceral adipocyte size was significantly smaller compared to subcutaneous (μ = −0·51, P = 1·57⋅10^−7^, *τ^2^* = 0·037, P*_τ_* = < 0·0001, *I^2^* = 86·2%) (Supplementary Figure 13), replicating the finding from our previous study (14). This increase in subcutaneous adipocyte size is particularly evident in the GTEX cohort compared to all other cohorts collectively (P_ALL_ = 3·71⋅10^−35^, β_ALL_ = 771·77) and individually (P_Leipzig_ = 2·30⋅10^−23^, β_Leipzig_ = 773·61; P_Munich_ = 1·19⋅10^−9^, β_Munich_ = 728·21; P_Hohenheim_ = 6·97⋅10^−24^, β_Hohenheim_ = 2300·19; P_Endox_ = 2·93⋅10^−24^, β_Endox_ = 1004·69). Taken together, these results provide evidence that age, sex and T2D status are associated with visceral adipocyte hypertrophy.

### Adipocyte hypertrophy is linked to clinical phenotypes in the context of cardiometabolic disease

To elucidate the relationship between adipocyte hypertrophy and cardiometabolic disease risk, we correlated mean adipocyte size measurements with relevant clinical biomarkers and adipokine levels utilizing data from the Leipzig cohort (Figure 2A-D).

Mean adipocyte size was positively correlated with levels of plasma glycated hemoglobin (mmol/mol) (P_SC_ = 1·06⋅10^−4^, β_SC_ = 2·02 and P_VC_ = 1·04⋅10^−3^, β_VC_ = 1·93) and with fasting plasma glucose in subcutaneous (P_SC_LOG10_ = 3·94⋅10^−5^, β_SC_LOG10_ = 0·013) and visceral adipose tissue (P_VC_LOG10_ = 5·06⋅10^−5^, β_VC_LOG10_ = 0·015). Furthermore, adiponectin was negatively (P_SC_LOG10_ = 0·011, β_SC_LOG10_ = −0·045, P_VC_LOG10_ = 0·017, β_VC_LOG10_ = −0·052) and leptin positively correlated with adipocyte size in both adipose depots (P_SC_LOG10_ = 2·03⋅10^−7^, β_SC_LOG10_ = 0·042, P_VC_LOG10_ = 3·19⋅10^−5^, β_VC_LOG10_ = 0·042). Lastly, the data revealed a positive association with interleukin 6 levels in visceral adipose tissue (P_VC_LOG10_ = 0·012, β_VC_LOG10_ = 0·14) (Figure 2A). These results are significant after controlling for FDR (Benjamini–Hochberg), (q-value = 0·036).

Next, we investigated the associations of adipocyte hypertrophy with blood lipid levels (Figure 2C-D). We found that mean adipocyte size correlated positively with levels of triglycerides (mmol/mol) (P_SC_LOG10_ = 3·53⋅10^−7^, β_SC_LOG10_ = 0·032 and P_VC_LOG10_ = 0·013, β_VC_LOG10_ = 0·019), and the triglycerides to HDL cholesterol ratio (P_SC_LOG10_ = 1·32⋅10^−9^, β_SC_LOG10_ = 0·049 and P_VC_LOG10_ = 5·15⋅10^−5^, β_VC_LOG10_ = 0·039). We further observed a negative correlation with HDL cholesterol (P_SC_ = 2·97⋅10^−7^, β_SC_ = −0·052 and P_VC_ = 8·28⋅10^−9^, β_VC_ = −0·069) (Figure 2D). These results were significant after controlling for FDR (Benjamini–Hochberg), (q-value = 0·03). Interestingly, we observed no significant difference in levels of LDL cholesterol (P_SC_ = 0·38, β_SC_ = 0·027 and P_VC_ = 0·31, β_VC_ = 0·036) and total cholesterol (P_SC_ = 0·22, β_SC_ = 0·040 and P_VC_ = 0·96, β_VC_ = 0·0020).

Lastly, sex-stratified analyses revealed no statistically significant differences in effect sizes between males and females (Supplementary Figures 14-15).

### GWAS on adipocyte size reveals novel genetic associations with adipocyte hypertrophy

To identify genetic determinants of adipocyte size and adipocyte hypertrophy, we performed the largest sex-combined and sex-stratified GWAS of adipocyte morphology to date, analyzing five independent cohorts of European ancestry in both subcutaneous (N=2,066) and visceral (N=1,878) adipose tissues. Quantile-quantile plots showed no evidence of systematic inflation across our GWAS for all five adipocyte size phenotypes (Supplementary Figure 16). We identified four depot-dependent genetic signals reaching genome-wide significance (P < 5·0⋅10^−8^), represented by independent (r2 < 0·05) index variants (Supplementary Figure 17).

Among the genome-wide significant loci, we detected an association for the intronic variant *rs73184721* (chr3:175280793, MAF=6·54% in EUR) in the *NAALADL2* gene with an increased 95%-quantile of adipocyte size in visceral adipose tissue (β = 930·18, β_SD_ = 0·42, P = 7.01⋅10^−9^) (Figure 3A). The association remained after adjusting for BMI (β_SD_ = 0·27, P = 1.19⋅10^−4^) and did not show sex heterogeneity (P = 0·81). Furthermore, the variant was also associated with increased mean adipocyte size in visceral adipose tissue (β_SD_ = 0·31, P = 3.87⋅10^−5^).

**Figure 3.**
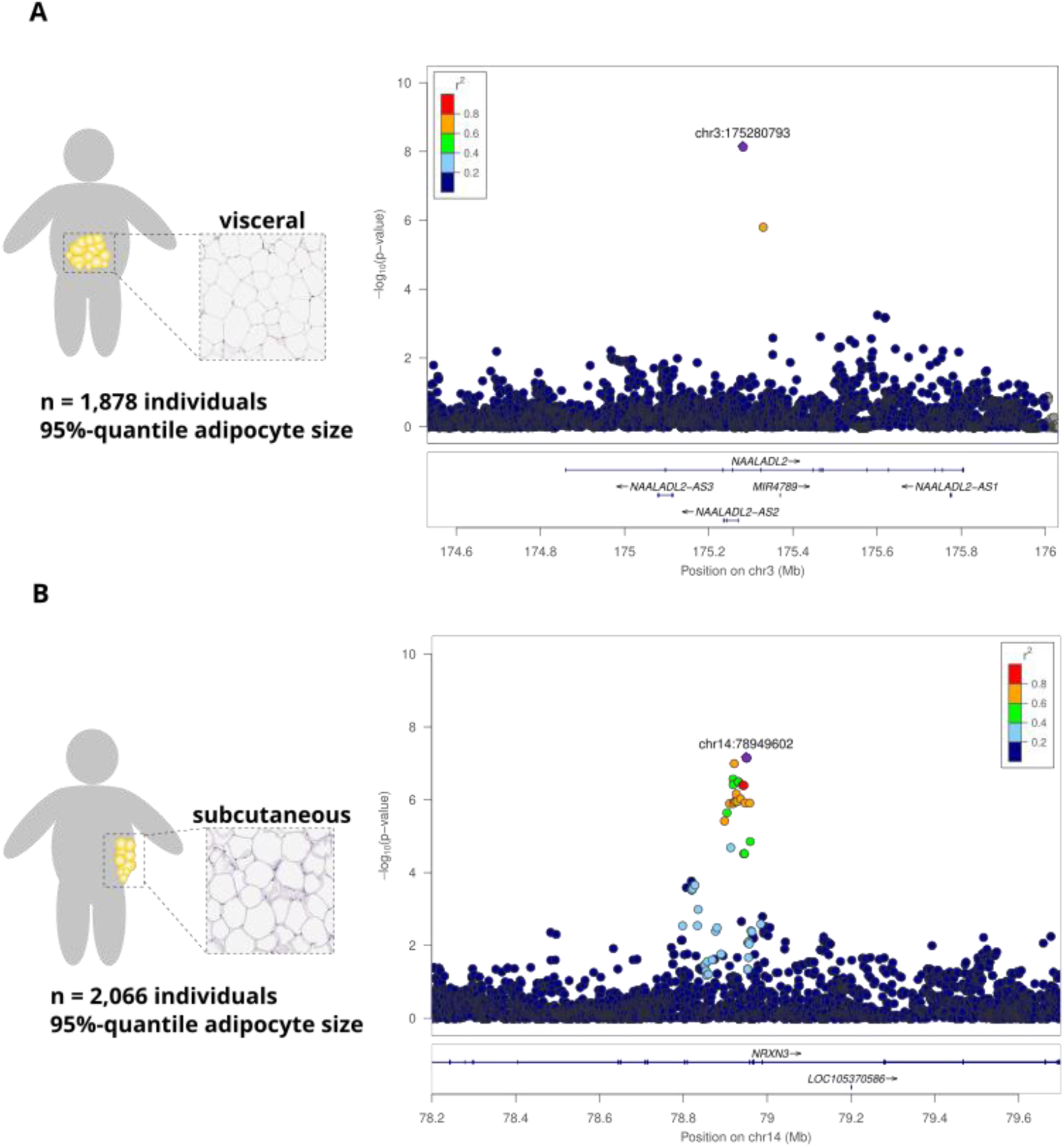
**A)** LocusZoom plot of rs73184721 (chr3:175280793) and its association with the 95%-quantile of adipocyte size in visceral adipose tissue. **B)** LocusZoom plot of rs200047724 (chr14:78949602) and its association with the 95%-quantile of adipocyte size in subcutaneous adipose tissue. For both plots, a region of +/− 750 kb has been plotted around the variant. The colour of the dots indicates the degree of LD between the lead variant and the other variants, the R2 values are based on the GTEX sequence data.

Additionally, we detected an association between the small indel intronic variant *rs200047724* (chr14:78949603, MAF=2.14% in EUR), located in the *NRXN3* gene, and an increased 95%-quantile of adipocyte size in subcutaneous adipose tissue in our meta-analysis across all five cohorts (β = 1592.92, β_SD_ = 0·60, P = 6·72⋅10^−8^) (Figure 3B).

Sex-stratified analyses revealed genetic determinants specific to females (Supplementary Figure 18-19), but no genetic determinants specific to males (Supplementary Figure 20-21). We identified an association between the intergenic variant *rs140503338* (chr14:68826371, MAF=2.15% in EUR) and increased mean adipocyte size exclusively in females in subcutaneous adipose tissue meta-analyzing all five cohorts (β = 956·06, β_SD_ = 0·85, P = 2.70⋅10^−8^) (Supplementary Figure 22). This variant was also associated with an increased 95%-quantile of adipocyte size in females in subcutaneous adipose tissue (β_SD_ = 0·78, P = 2.86⋅10^−7^) and remained significant after adjusting for BMI (β_SD_ = 0·66, P = 4·78⋅10^−6^). While the genetic association of *rs14053338* with adipocyte size and hypertrophy in females was specific to subcutaneous adipose tissue, we observed a nominal association in the meta-analysis for mean visceral adipocyte size (β_SD_ = 0·32, P = 0·044) and 95%-quantile of visceral adipocyte size (β_SD_ = 0·35, P = 0·028) in females.

Finally, we identified an association between the intronic variant *rs11656704* (chr17:19789496, MAF=22.0%) in the *ULK2* gene with a decreased 95%-quantile of visceral adipocyte size in females (β = −700·64, β_SD_ = −0·31, P = 4·50⋅10^−8^). This signal remained significant after adjusting for BMI (β_SD_ = −0·26, P = 7.36⋅10^−7^) and mean adipocyte size (β_SD_ = −0·29, P = 4·62⋅10^−7^).

To evaluate the clinical relevance of these genetic associations in the context of metabolic disease, we performed phenome-wide association studies (PheWAS). Interestingly, the female-specific variant *rs14053338* was significantly associated with plasma glycated hemoglobin levels (β_SD_ = 0·65, P = 0·0081) (Table 2) and nominally associated with BMI (β_SD_ = 0·42, P = 0·016) and 2-hour plasma glucose (β_SD_ = 1.25, P = 0·046). The other female-specific variant in *ULK2* was associated with WHR (β_SD_ = −0·54, P = 0·0056) and plasma glycated hemoglobin levels (β_SD_ = −0·26, P = 0·0055) in our PheWAS (Table 2).

**Table 2.** PheWAS results for the three lead variants. P-value and effect size from the inverse normal quantile transformed traits are reported. These analyses are run in the Leipzig Cohort. All values with a P-value below 0·05 (nominal significance) have been highlighted in bold.

| Trait | rs73184721 (chr3)<br>(P / $\beta_{SD}$) | rs200047724<br>(chr14) (P / $\beta_{SD}$) | rs140503338<br>(chr14) (P / $\beta_{SD}$)<br>(Females only) | rs11656704<br>(chr17) (P / $\beta_{SD}$)<br>(Females only) |
| --- | --- | --- | --- | --- |
| WHR | **0.012 / -0.39** | 0.054 / -0.58 | 0.27 / 0.58 | **0.0056 / -0.54** |
| BMI | 0.70 / -0.027 | 0.12 / -0.16 | **0.016 / 0.42** | 0.27 / -0.075 |
| plasma glycated hemoglobin (%) | 0.58 / 0.070 | 0.057 / -0.34 | **0.0081 / 0.65** | **0.0055 / -0.26** |
| fasting plasma glucose | 0.30 / -0.11 | 0.72 / -0.056 | 0.062 / 0.38 | 0.060 / -0.14 |
| 2h plasma glucose | 0.95 / 0.020 | 0.063 / 1.05 | **0.046 / 1.25** | 0.60 / -0.15 |
| fasting plasma insulin | 0.41 / -0.12 | 0.49 / -0.18 | 0.35 / 0.27 | 0.16 / -0.15 |
| Interleukin-6 | 0.88 / -0.066 | 0.59 / 0.28 | 0.89 / -0.072 | 0.32 / -0.21 |
| Adiponectin | 0.79 / -0.10 | 0.84 / -0.11 | 0.071 / -1.16 | 0.12 / 0.40 |
| C-reactive protein | 0.62 / -0.050 | 0.94 / 0.012 | 0.17 / 0.26 | 0.52 / -0.048 |

## Discussion

Our novel approach of WSI segmentation enables comprehensive detection and quantification of adipocytes, including very large hypertrophic cells missed by previous methods. Our approach allows us to better characterize the metabolic impact of adipocyte hypertrophy and to investigate its genetic architecture at an unprecedented scale.

We show that increased adipocyte size is associated with an adverse metabolic profile, consistent with previously reported elevated risk of type 2 diabetes and increased plasma fasting glucose (8). This relationship with fasting plasma glucose appears stronger in visceral compared to subcutaneous adipose tissue (β_SC_LOG10_ = 0·013 and β_VC_LOG10_ = 0·015), in line with the established metabolic significance of visceral adiposity (8). The positive correlation between adipocyte size and levels of glycated hemoglobin corroborates findings from Keshavjee *et al.*(*29*), who reported that increased average adipose cell area in visceral adipose tissue was associated with persistently elevated glycated hemoglobin following bariatric surgery (post-Roux-en-Y gastric bypass).

The inverse relationship between adiponectin levels and mean adipocyte size is consistent with previous findings (8). Interestingly, we detected a significant association between interleukin-6 and mean adipocyte size specifically in visceral but not in subcutaneous adipose tissue. This is inconsistent with (30), where levels of interleukin 6 were higher in diabetic and non-diabetic obese patients compared to control subjects in subcutaneous adipose tissue (Figure 2). The increased mean size of adipocytes in men and with increased age in visceral adipose tissue is consistent with the meta-analysis in (8). However, the finding of no increase in the mean size of adipocytes with age in subcutaneous adipose tissue is not. Additionally, the decreased mean adipocyte size in visceral depots is also consistent with the literature (8) (Supplementary Figure 5-13). Importantly, these results demonstrate significant heterogeneity between cohorts, suggesting that the surgery approach, anatomical location of the biopsy site and the study population characteristics are potential modifiers of adipocyte size.

Notably, we did not detect a statistically significant increase in fasting plasma insulin levels with an increased mean size of adipocytes, contrary to previous findings (8) (Figure 2). We also observed no sex-specific differences in the metabolic impact of adipocyte hypertrophy. (Supplementary Figure 14-15).

Our study represents a substantial advance in understanding the genetic architecture of adipocyte morphology. Only two previous studies, including our earlier work, have investigated this area (14,17), both failing to identify genome-wide significant associations (P < 5.0⋅10^−8^). We report four novel genetic loci associated with adipocyte size phenotypes. The variant rs73184721 is an intronic variant in the large *NAALADL2* gene, whose function remains largely unknown. However, another gene close to the associated variant is *NLGN1*, which acts as a splice site-specific ligand for β-neurexins. Intriguingly, this variant has been nominally associated with increased pancreas fat percentage and increased subcutaneous adipose tissue volume (31). *NLGN1* has also been linked to predicted visceral adipose tissue mass (12), BMI (32), and type 2 diabetes (33), suggesting that the effect on visceral adipocyte hypertrophy might be mediated through *NLGN1* expression, though this merits further investigation.

The association of rs200047724 in the *NRXN3* gene with the increased 95%-quantile of adipocyte size is particularly intriguing. *NRXN3*, neurexin 3, encodes proteins functioning as receptors and cell adhesion molecules primarily expressed in the brain. Intriguingly, the *NRXN3* gene has previously been associated with BMI (34), waist circumference (35), and altered methylation expression in extremely obese individuals (36), suggesting a neuronal component in the regulation of adipocyte morphology.

The female-specific variant rs140503338 is located near a gene cluster containing *RAD51B,* a member of the *RAD51* protein family essential for DNA repair by homologous recombination. This gene has been previously associated with waist-hip ratio (both BMI-adjusted (37) and unadjusted (38), and is most highly expressed in the uterus, potentially explaining its sex-specific effects (39). We therefore speculate that the observed association with adipocyte size might be mediated through the *RAD51B* gene, though this requires further investigation.

The other female-specific variant, rs11656704, is an intronic variant in the gene *ULK2* located near *AKAP10,* which has been previously associated with BMI (40). While this variant showed no association with BMI in our analysis, it was significantly associated with decreased waist-hip ratio and reduced plasma glycated hemoglobin levels (Table 2). This suggests that *AKAP10* may influence adipocyte morphology through mechanisms independent of overall adiposity, though this is speculative and the involvement of *AKAP10* as a causal effector in adipocyte morphology warrants further investigation.

Given the unusual linkage disequilibrium patterns observed for rs73184721 and rs140503338, we verified these patterns against both the 1000 Genomes (43) and TOPMed (44) reference panels, confirming their validity (41).

Interestingly, our colocalisation analysis revealed shared genetic architecture between adipocyte size and HDL cholesterol levels, with effects in the same direction as observed in our epidemiological analyses—increased visceral adipocyte size corresponded with decreased HDL cholesterol (Figure 2D). While the top variant (rs2070512) is an expression quantitative trait locus (eQTL) for *CCDC116* in both adipose depots and for *UBE2L3* and *YDJC* in subcutaneous adipose tissue, none of these genes have established connections to adipose biology, suggesting potential novel pathways linking adipocyte morphology to lipid metabolism.

To our knowledge, this study is the first to report genome-wide significant genetic associations with adipocyte size, providing important insights into the genetic architecture underlying this cellular phenotype. The identification of both sex-combined and female-specific loci offers intriguing insights into the sexual dimorphism of fat distribution and metabolic risk, a phenomenon long observed clinically but poorly understood mechanistically.

Intriguingly, two of our significant loci point toward a potential brain-adipose signaling axis. The association of *NRXN3* with subcutaneous adipocyte morphology suggests a connection between neuronal signaling and peripheral adipocyte behavior. Similarly, the proximity of *NLGN1* to our lead variant rs73184721 further strengthens this hypothesis, as neuroligins and neurexins form transynaptic complexes crucial for synaptic function. This convergence of neuronal adhesion molecules in adipocyte size regulation suggests previously unexplored central nervous system influences on adipocyte biology, potentially coordinating energy homeostasis between brain and adipose tissue.

We stress that due to the lack of comparable genetic studies in this field, direct replication of our findings is currently not possible. However, as computational methods continue to advance, the combination of genetic data with deep learning-based histological analysis will likely yield further insights into the cellular foundations of metabolic disease and provide further validation. Our findings highlight the adipocyte as a genetically regulated and responsive cell type whose morphological characteristics play an important role in the link between obesity and metabolic complications.

## Data Availability

All data produced in the present study are available upon reasonable request to the authors

## Availability and implementation

The method is implemented in Python, making use of the PyTorch library. The code for training the model can be found at https://github.com/Nellaker-group/PyTorchUnet, and the code for running predictions with the model can be found here https://github.com/Nellaker-group/UNetHAPPY.

## Acknowledgments

We would like to thank the Core Facility for Comparative Experimental Pathology at Klinikum Rechts der Isar, Technical University of Munich for their technical support in obtaining the whole slide images.

## Author contributions

E.J., P.K., J.H., C.N., C.M.L., H.H. and M.C. conceived the study and designed the experiments. E.J., P.D.C. and C.N. wrote the code. E.J. and J.H. created ground truth annotations. E.J. performed the experimental analysis. J.H. and P.K. prepared and processed the adipose tissue histology samples. H.H., M.B., M.R, A.F., D.M, P.K. and M.C. collected the adipose tissue biopsies. E.J. conducted the genetic association studies of adipocyte morphology with input from T.H. and A.K. E.J. performed the quality control and imputation of the genetic data. E.J. and P.K. conducted the epidemiological studies of adipocyte morphology with input from P.D.C., C.N., and M.C.. E.J. and P.K. prepared the manuscript with input, revisions, and approval from M.C., C.A.G., H.H., M.B., T.H., C.M.L. and A.K. The supervision of the research was done by P.D.C., C.M.L., C.N., H.H. and M.C.

