## Supplementary Material for "Scalable Deep Learning of Histology Images Reveals Genetic and Phenotypic Determinants of Adipocyte Hypertrophy"

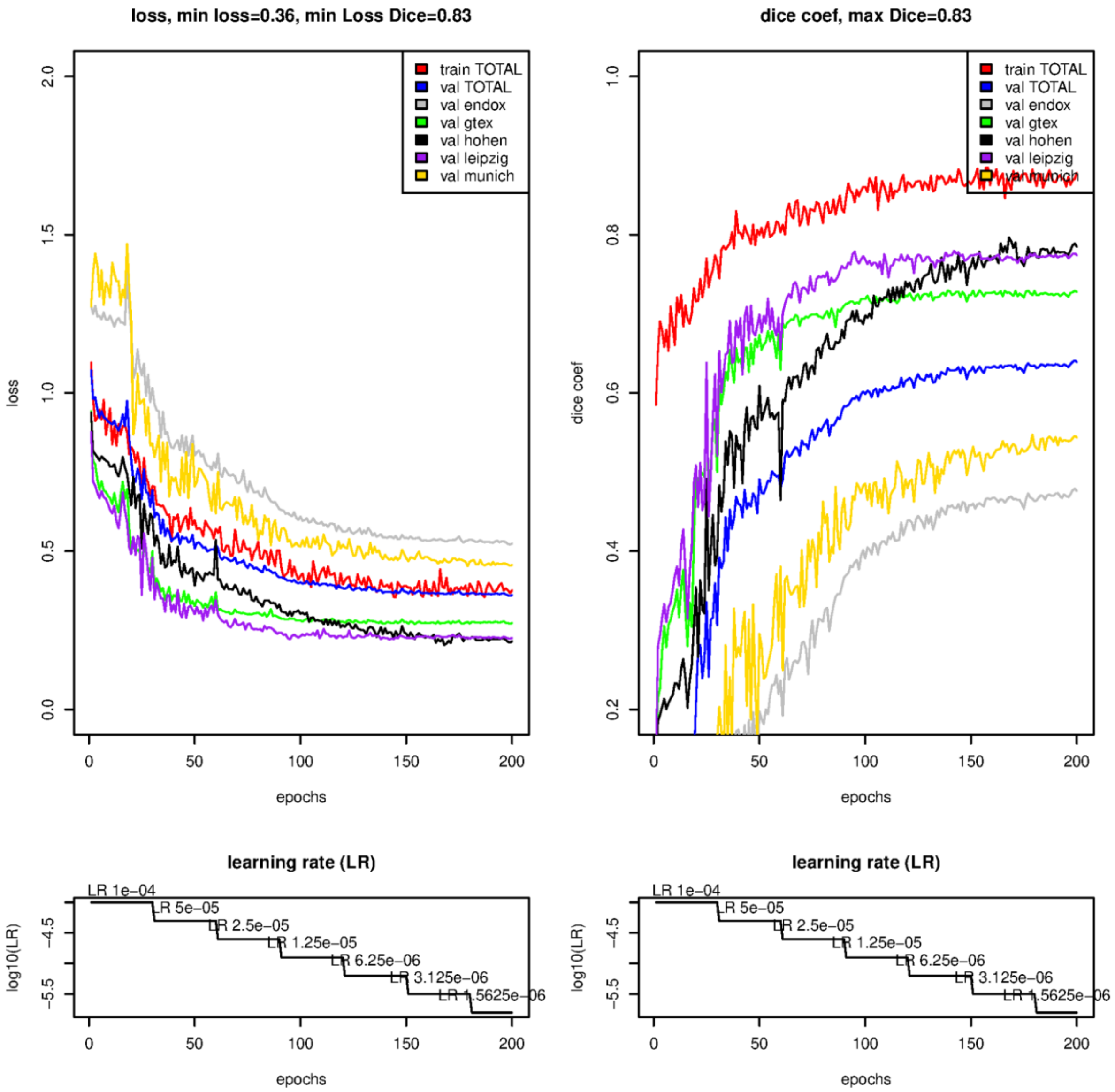


**Supplementary Figure 1.** Plot of loss per epoch for the training of our U-net model. In the left plot we show the total loss for training and validation (stratified on cohort). In the right plot we show the dice coefficient for training and validation (stratified on cohort). The lower plots show the learning rate used for training.

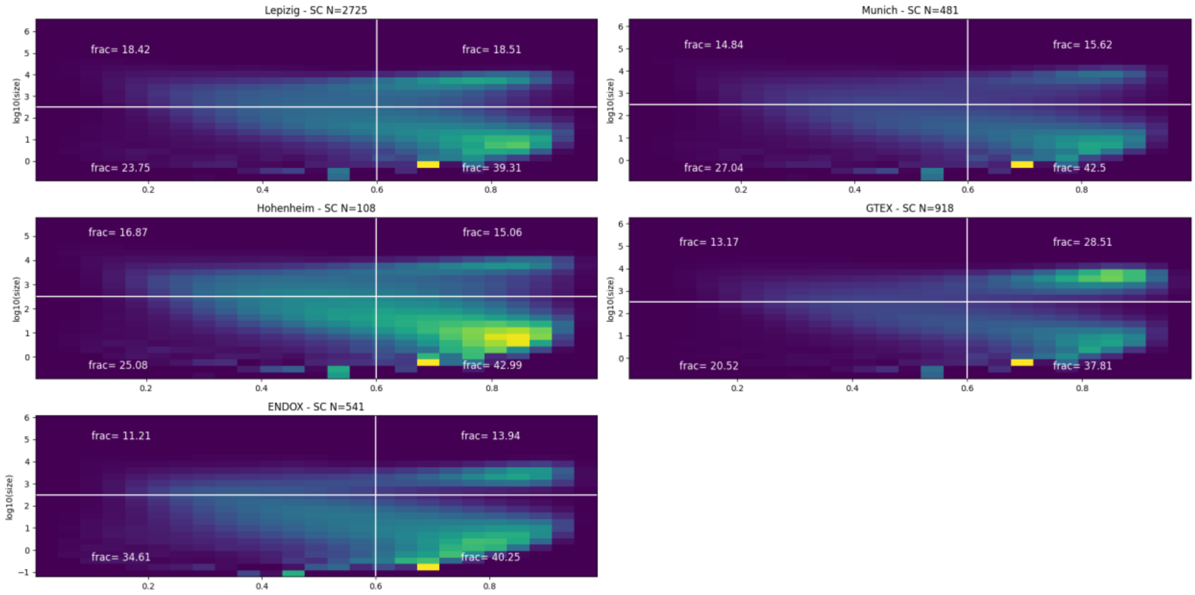


**Supplementary Figure 2.** 2D histogram of log10 of adipocyte size in subcutaneous adipose tissue (y-axis) and Polsby-popper roundness (x-axis). The white lines denote the cutoff used for filtering of adipocytes, the upper right rectangle are the adipocytes kept for further analysis. There is a plot for each cohort.

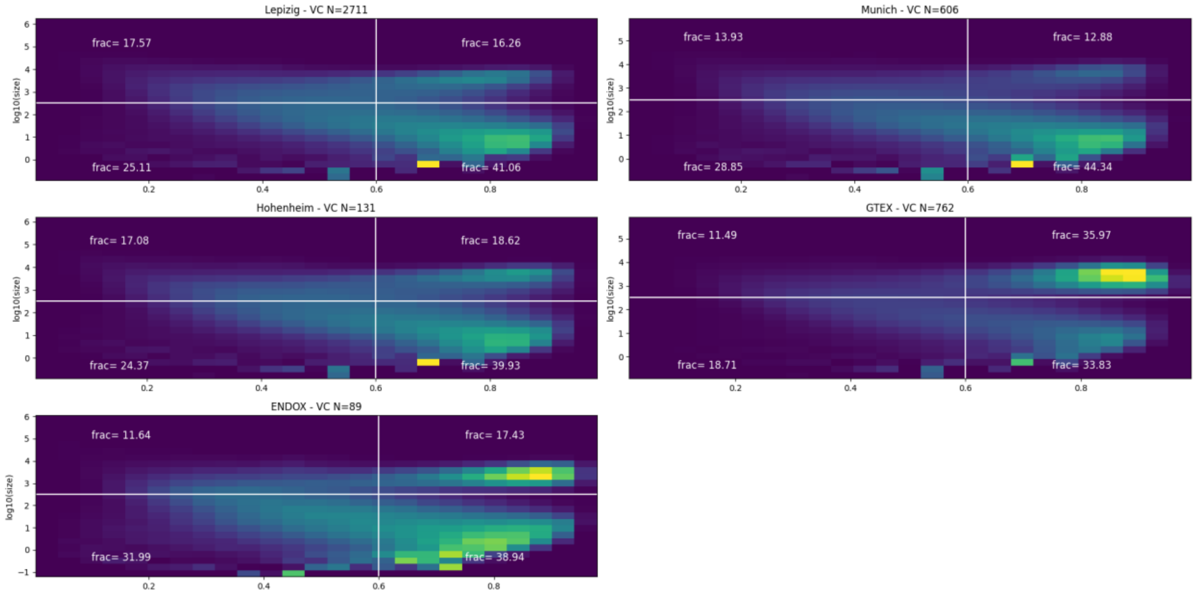


**Supplementary Figure 3.** The same as Supplementary Figure 2 however with visceral adipose tissue.

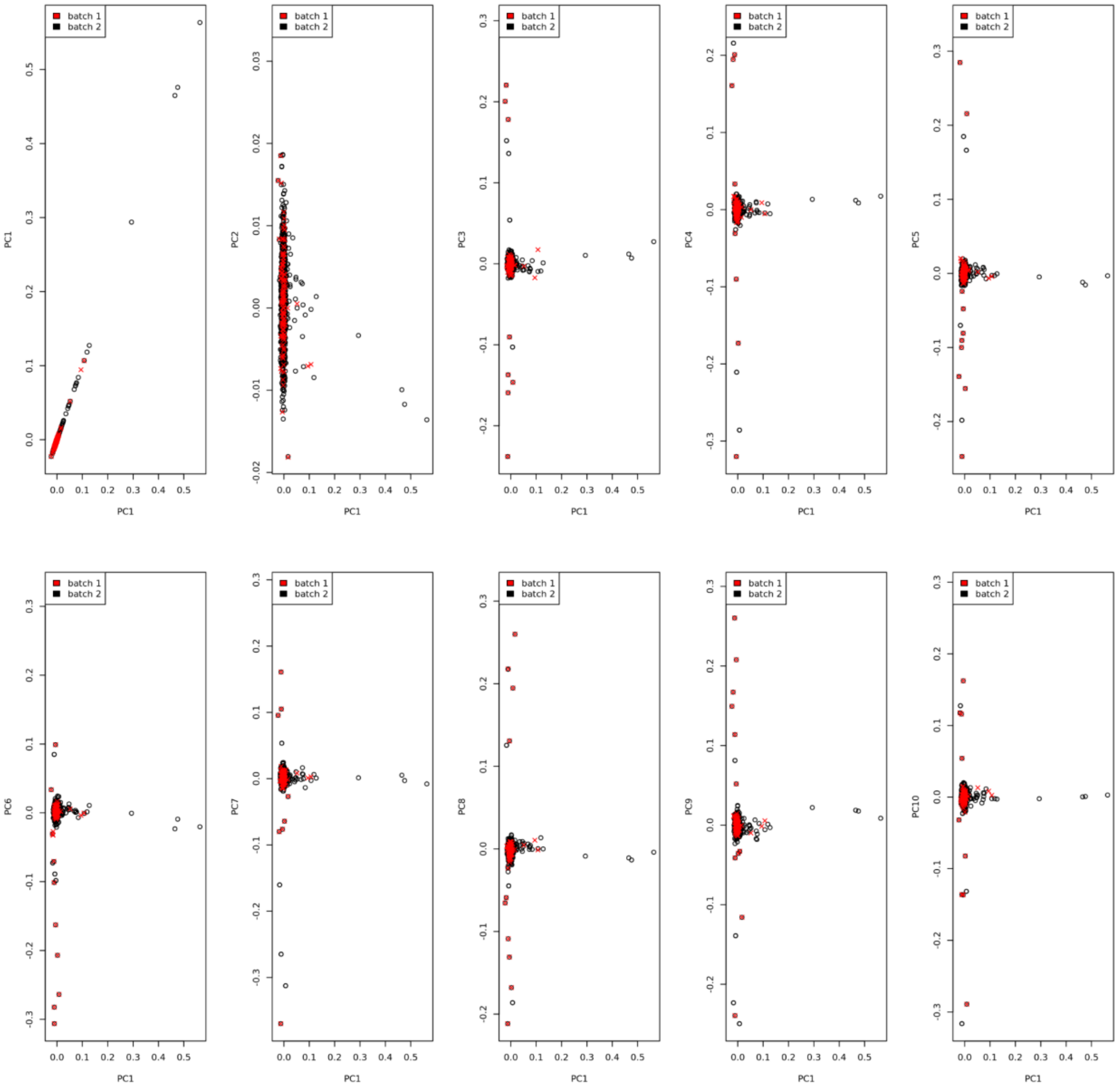


**Supplementary Figure 4.** Genetic principal component 1 plotted against principal components 1-10, of the two batches of the SNP-chip used for the Leipzig cohort. Each dot is coloured by which genotyping batch it is from.

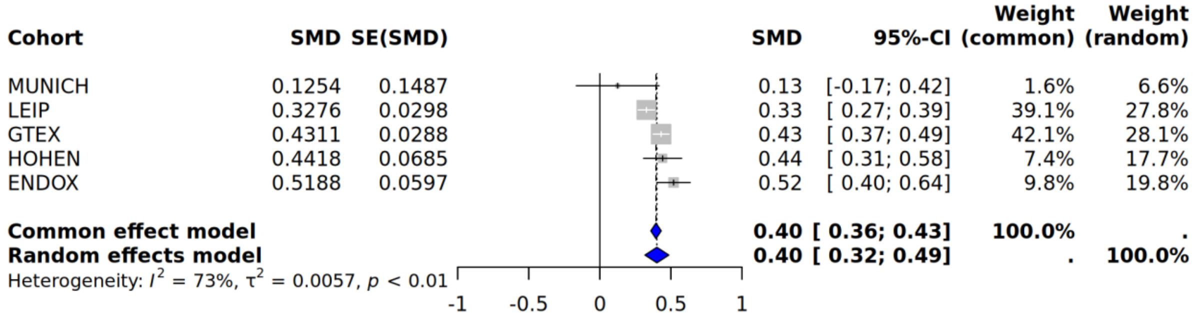


**Supplementary Figure 5.** Meta analysis of an association between BMI across cohorts and mean size of adipocytes in subcutaneous adipose tissue.

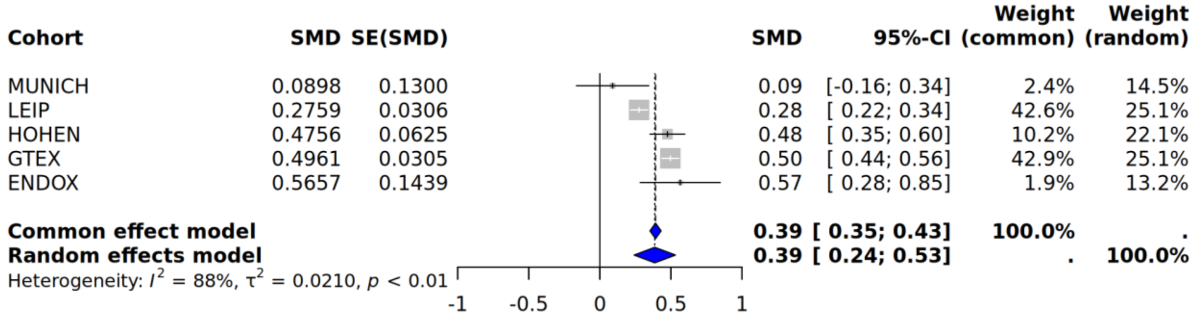


**Supplementary Figure 6.** Meta analysis of an association between BMI across cohorts and mean size of adipocytes in visceral adipose tissue.

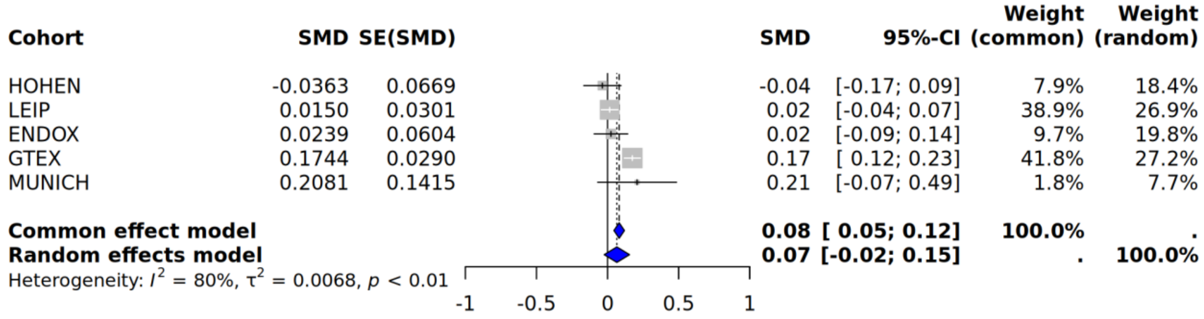


**Supplementary Figure 7.** Meta analysis of an association between age across cohorts and mean size of adipocytes in subcutaneous adipose tissue.

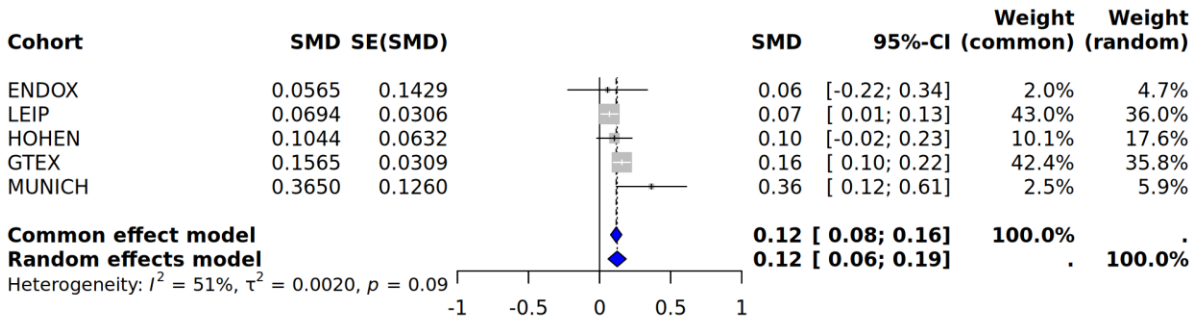


**Supplementary Figure 8.** Meta analysis of an association between age across cohorts and mean size of adipocytes in visceral adipose tissue.

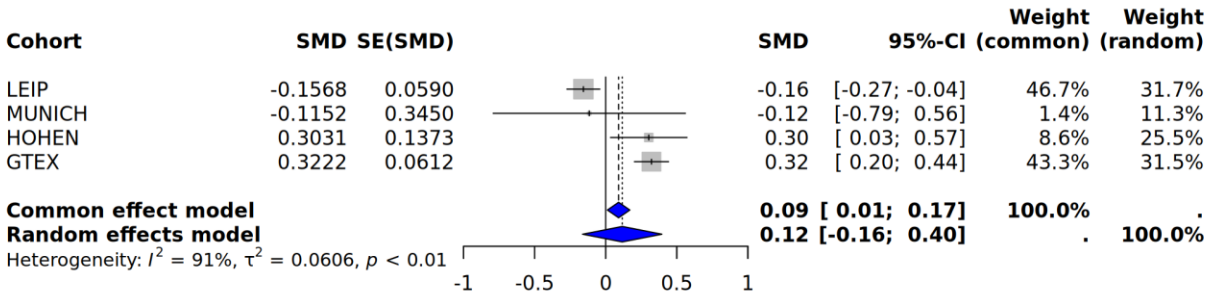


**Supplementary Figure 9.** Meta analysis of an association between sex across cohorts and mean size of adipocytes in subcutaneous adipose tissue.

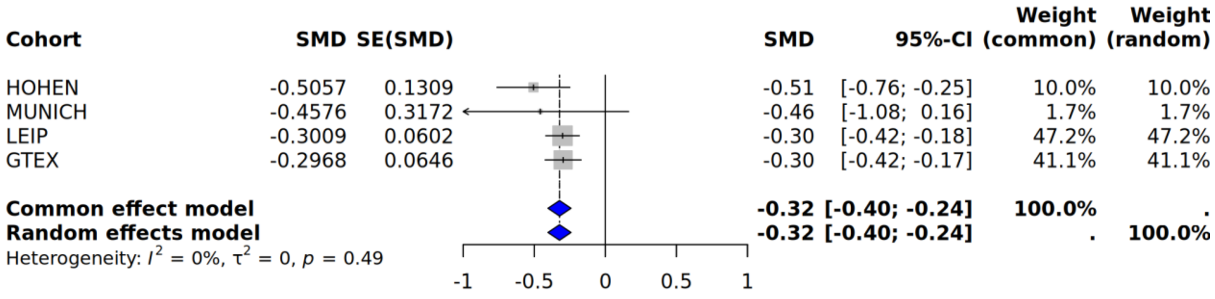

**Supplementary Figure 10.** Meta analysis of an association between sex across cohorts and mean size of adipocytes in visceral adipose tissue.

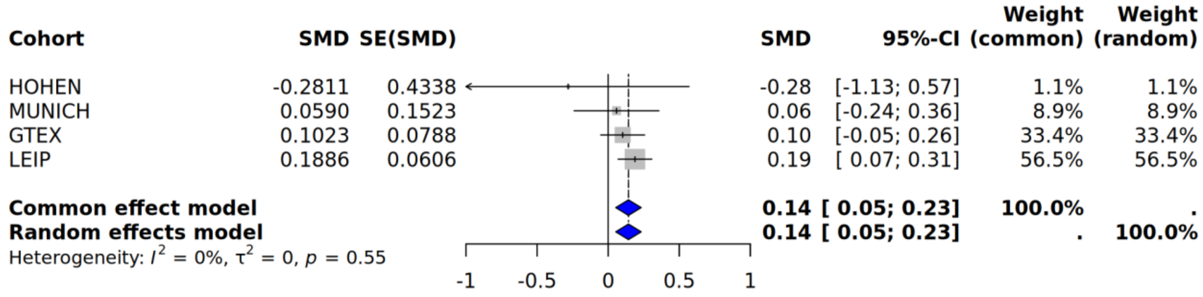


**Supplementary Figure 11.** Meta analysis of an association between type 2 diabetes status across cohorts and mean size of adipocytes in subcutaneous adipose tissue.

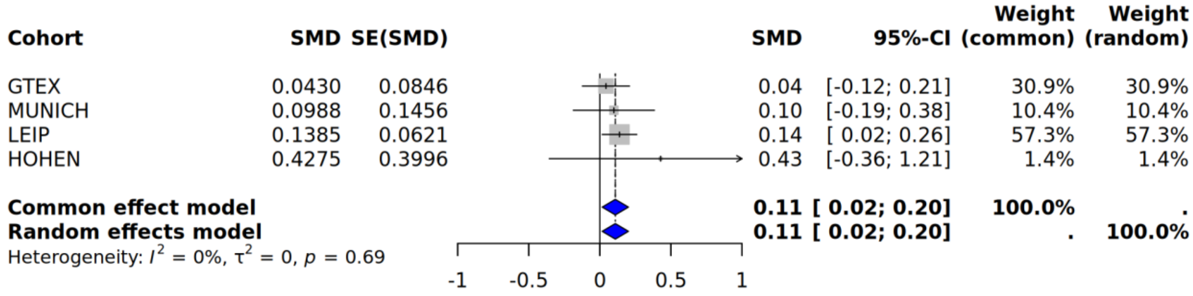


**Supplementary Figure 12.** Meta analysis of an association between type 2 diabetes status across cohorts and mean size of adipocytes in visceral adipose tissue.

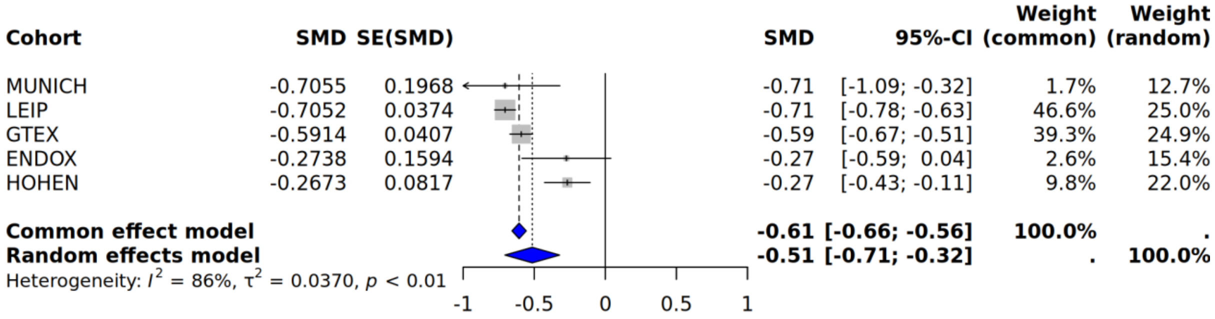

**Supplementary Figure 13.** Meta analysis of an association between depot (if visceral or subcutaneous) across cohorts and mean size of adipocytes.

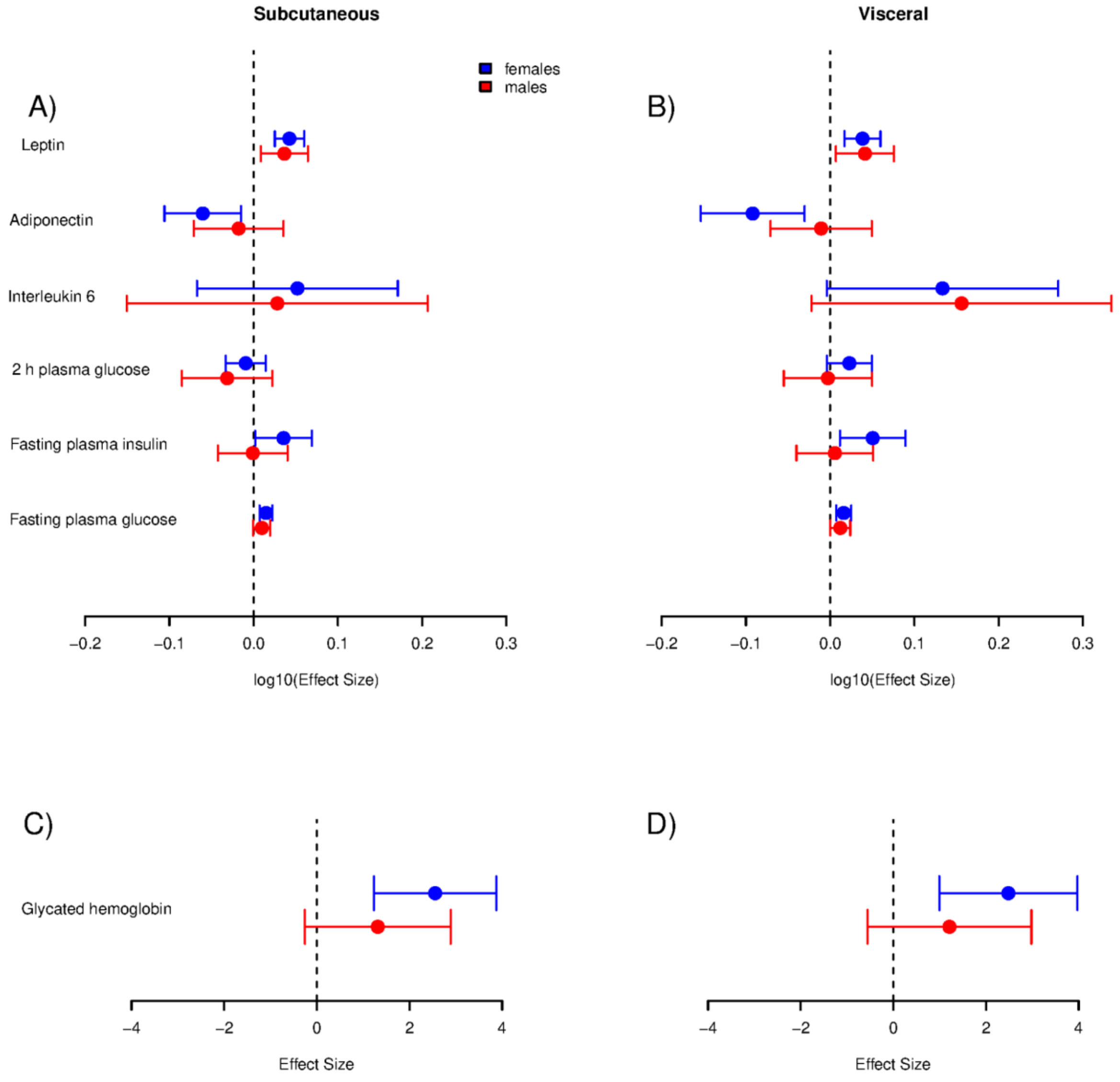


**Supplementary Figure 14.** Effect size of mean size of adipocytes on the listed glucose-homeostasis and inflammation related phenotypes, stratified by sex, in subcutaneous adipose tissue A) and C) and visceral adipose tissue B) and D) with the 95% confidence intervals.There are no statistically significant differences in effect size between males and females.

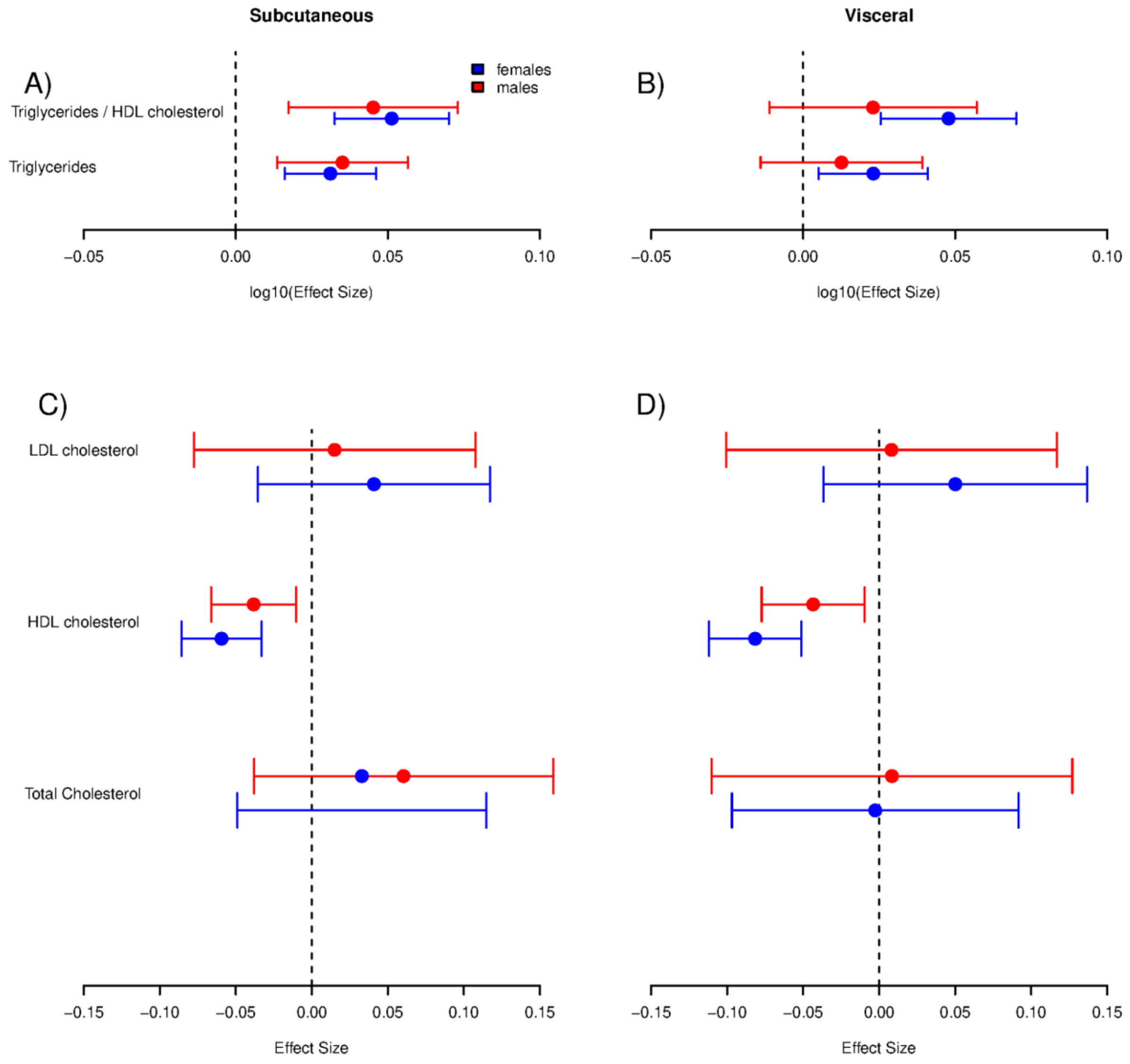


**Supplementary Figure 15.** Effect size of mean size of adipocytes on the listed cardiovascular related phenotypes, stratified by sex, in subcutaneous adipose tissue A) and C) and visceral adipose tissue B) and D) with the 95% confidence intervals. There are no statistically significant differences in effect size between males and females.

### Manhattan and QQ-plots of GWASes - combined

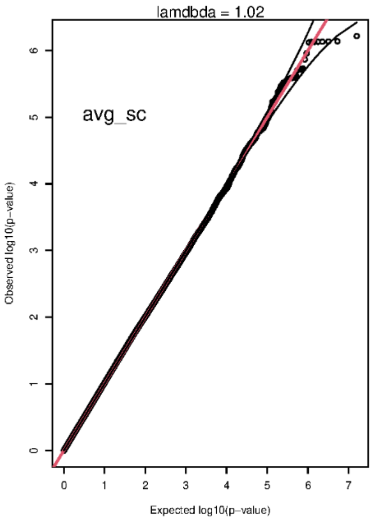


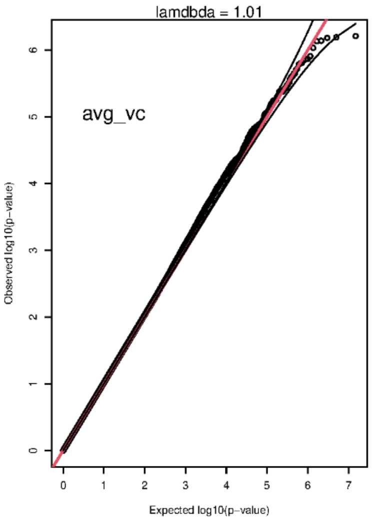


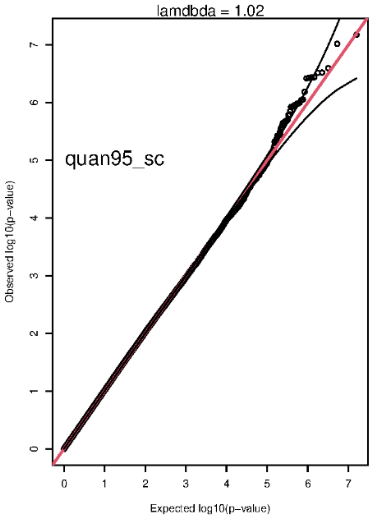


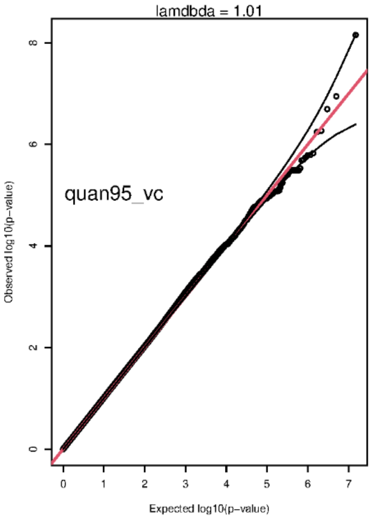


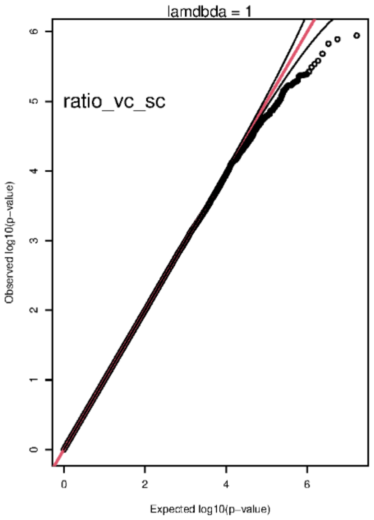

**Supplementary Figure 16.** QQ-plot for mean size of adipocytes in subcutaneous and visceral adipose tissue, 95%-quantile of adipocyte size in subcutaneous and visceral adipose tissue, and ratio of mean size of adipocytes between visceral and subcutaneous adipose tissue. Only variants present in all five cohorts are included.

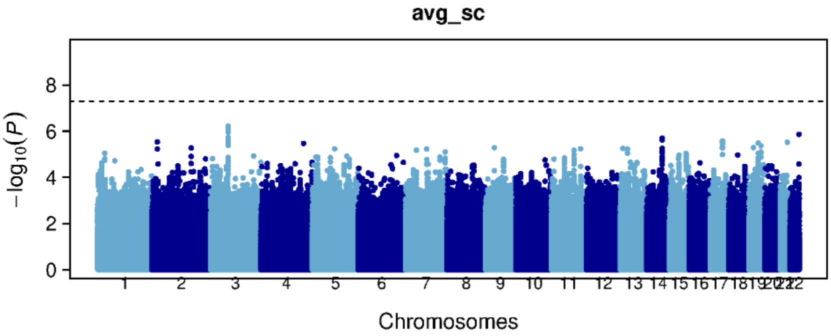
avg_sc
−log10(P)
Chromosomes

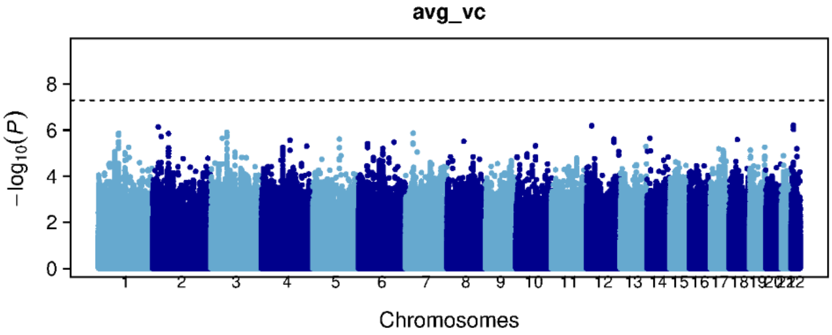
avg_vc
−log10(P)
Chromosomes

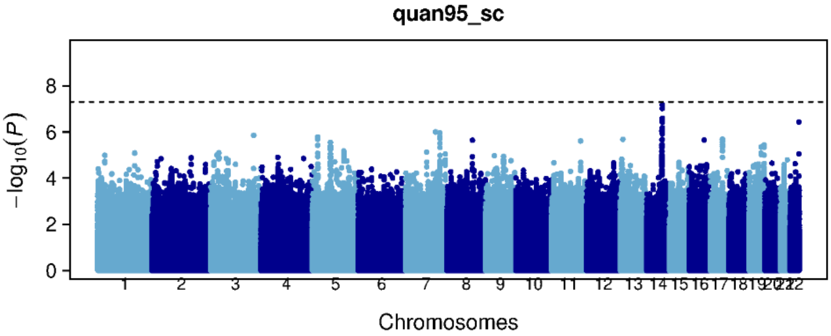
quan95_sc
−log10(P)
Chromosomes

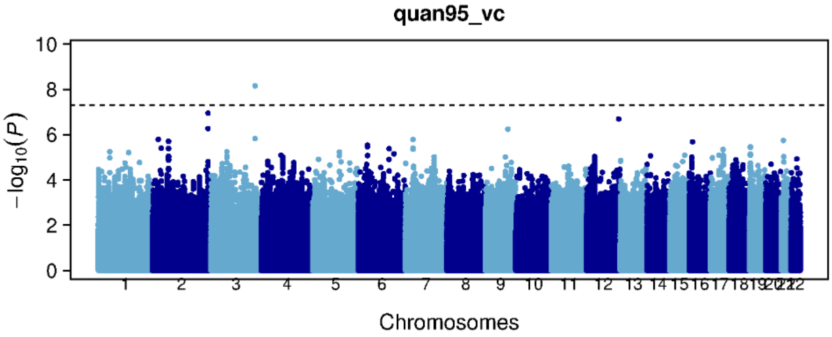
quan95_vc
−log10(P)
Chromosomes

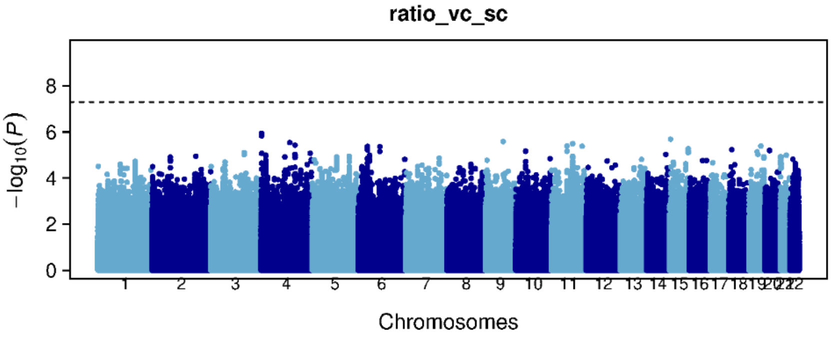
ratio_vc_sc
−log10(P)
Chromosomes

**Supplementary Figure 17.** Manhattan plot for mean size of adipocytes in subcutaneous and visceral adipose tissue, 95%-quantile of adipocyte size in subcutaneous and visceral adipose tissue, and ratio of mean size of adipocytes between visceral and subcutaneous adipose tissue. Only variants present in all five cohorts are included.

### Manhattan and QQ-plots of GWASes - females only

avg_sc

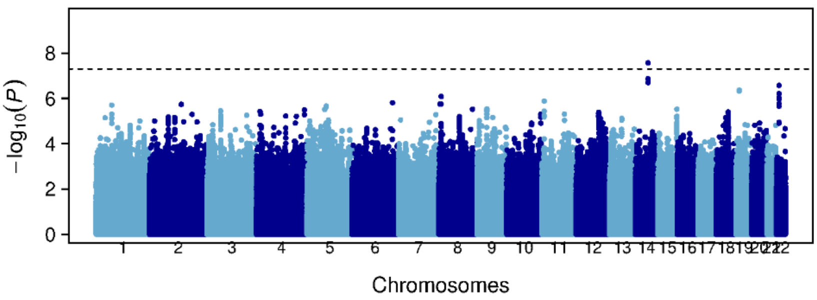


avg_vc

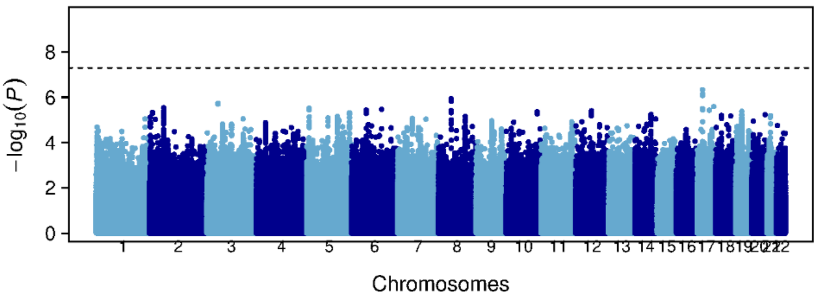


quan95_sc

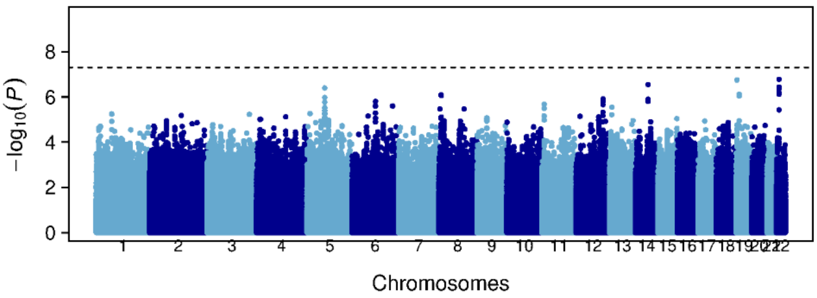


quan95_vc

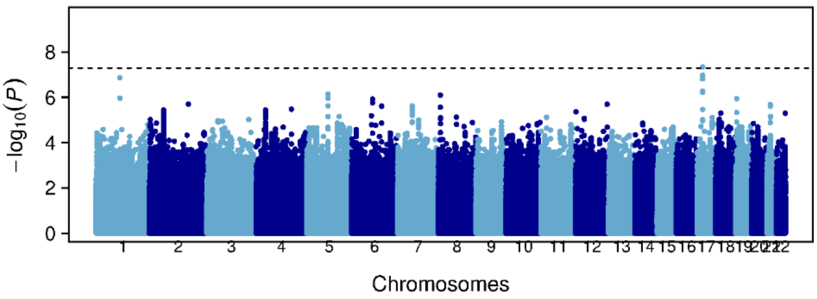


ratio_vc_sc

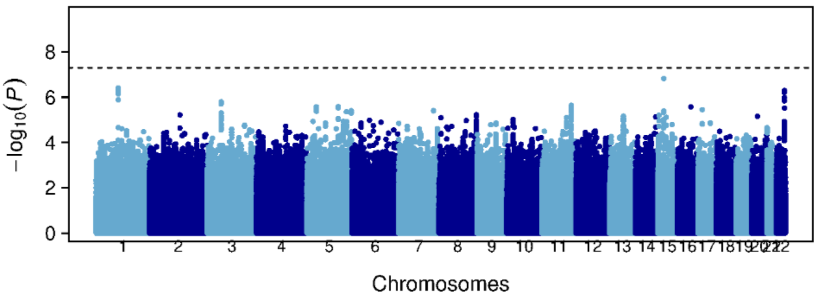

**Supplementary Figure 18.** Manhattan plot for mean size of adipocytes in subcutaneous and visceral adipose tissue, 95%-quantile of adipocyte size in subcutaneous and visceral adipose tissue, and ratio of mean size of adipocytes between visceral and subcutaneous adipose tissue. Only variants present in all five cohorts are included. Females only.

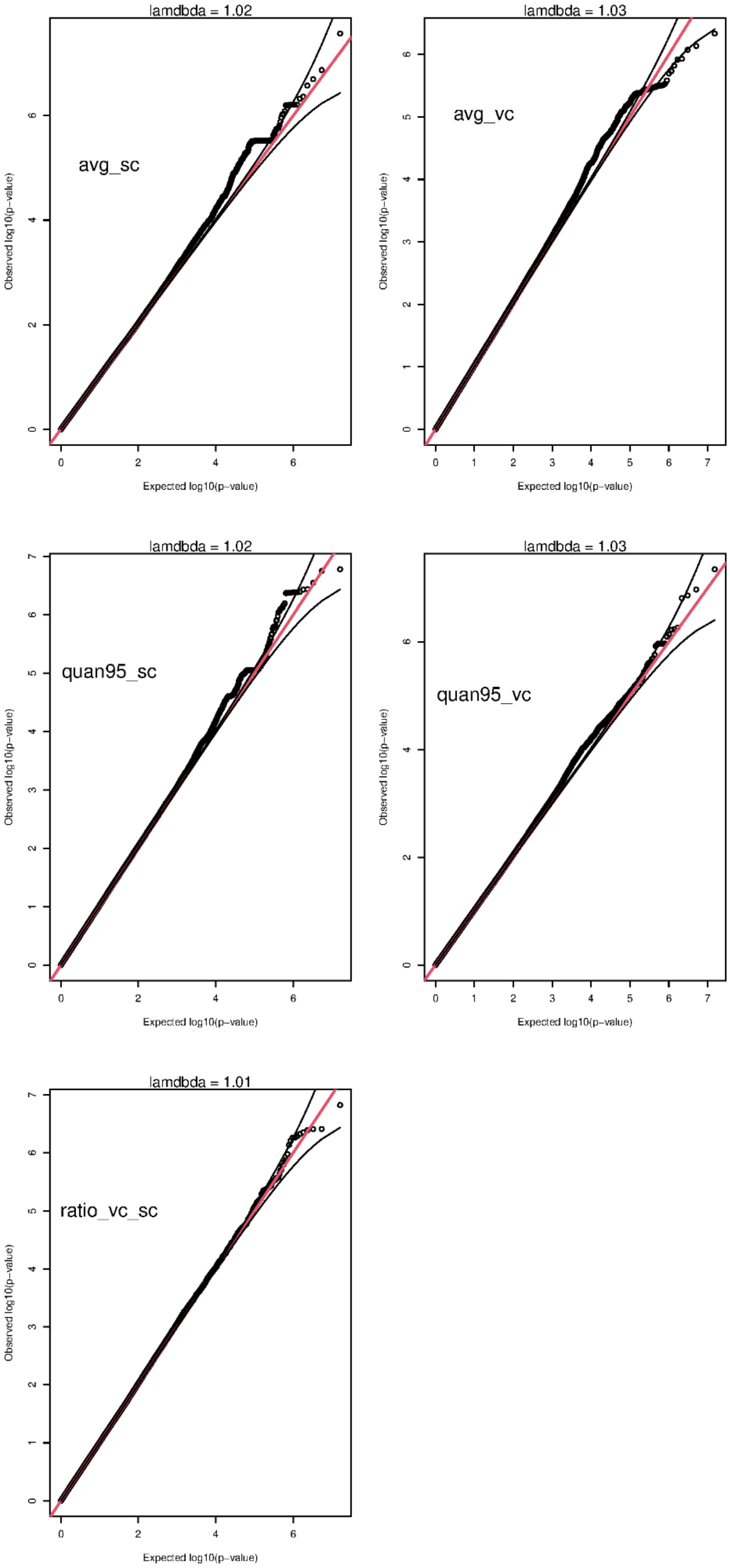

**Supplementary Figure 19.** QQ-plot for mean size of adipocytes in subcutaneous and visceral adipose tissue, 95%-quantile of adipocyte size in subcutaneous and visceral adipose tissue, and ratio of mean size of adipocytes between visceral and subcutaneous adipose tissue. Only variants present in all five cohorts are included. Females only.

### Manhattan and QQ-plots of GWASes - males only

**avg_sc**

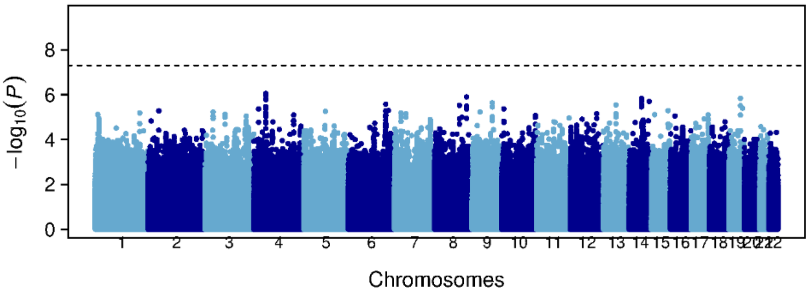


**avg_vc**

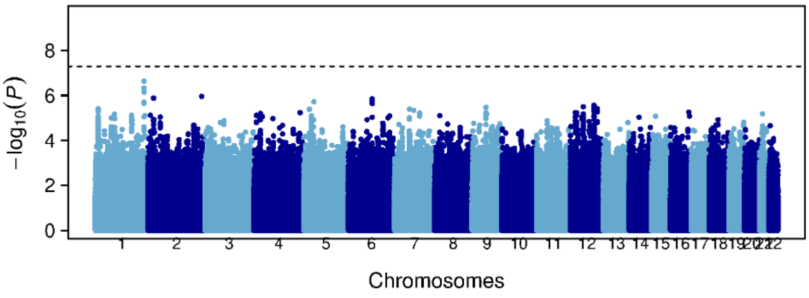


**quan95_sc**

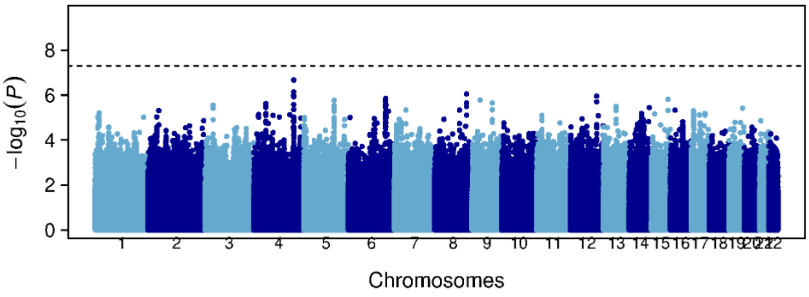


**quan95_vc**

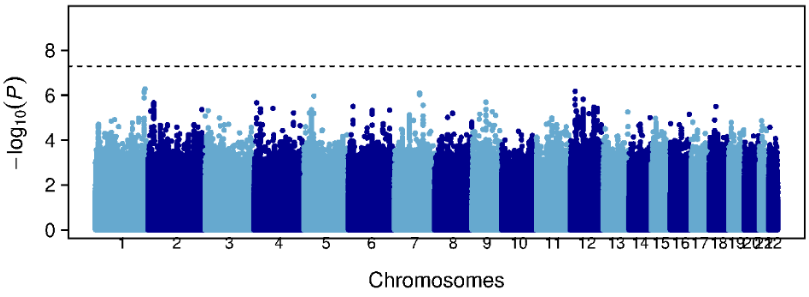


**ratio_vc_sc**

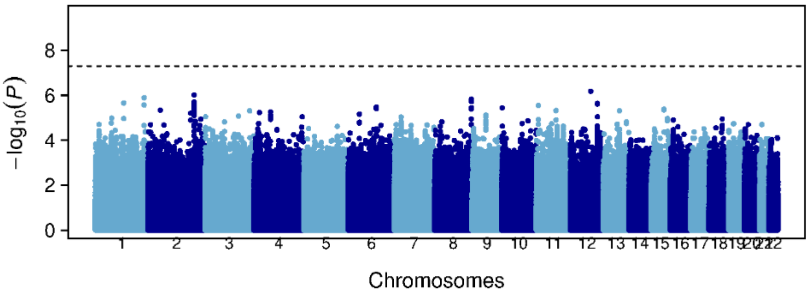

**Supplementary Figure 20.** Manhattan plot for mean size of adipocytes in subcutaneous and visceral adipose tissue, 95%-quantile of adipocyte size in subcutaneous and visceral adipose tissue, and ratio of mean size of adipocytes between visceral and subcutaneous adipose tissue. Only variants present in all five cohorts are included. Males only.

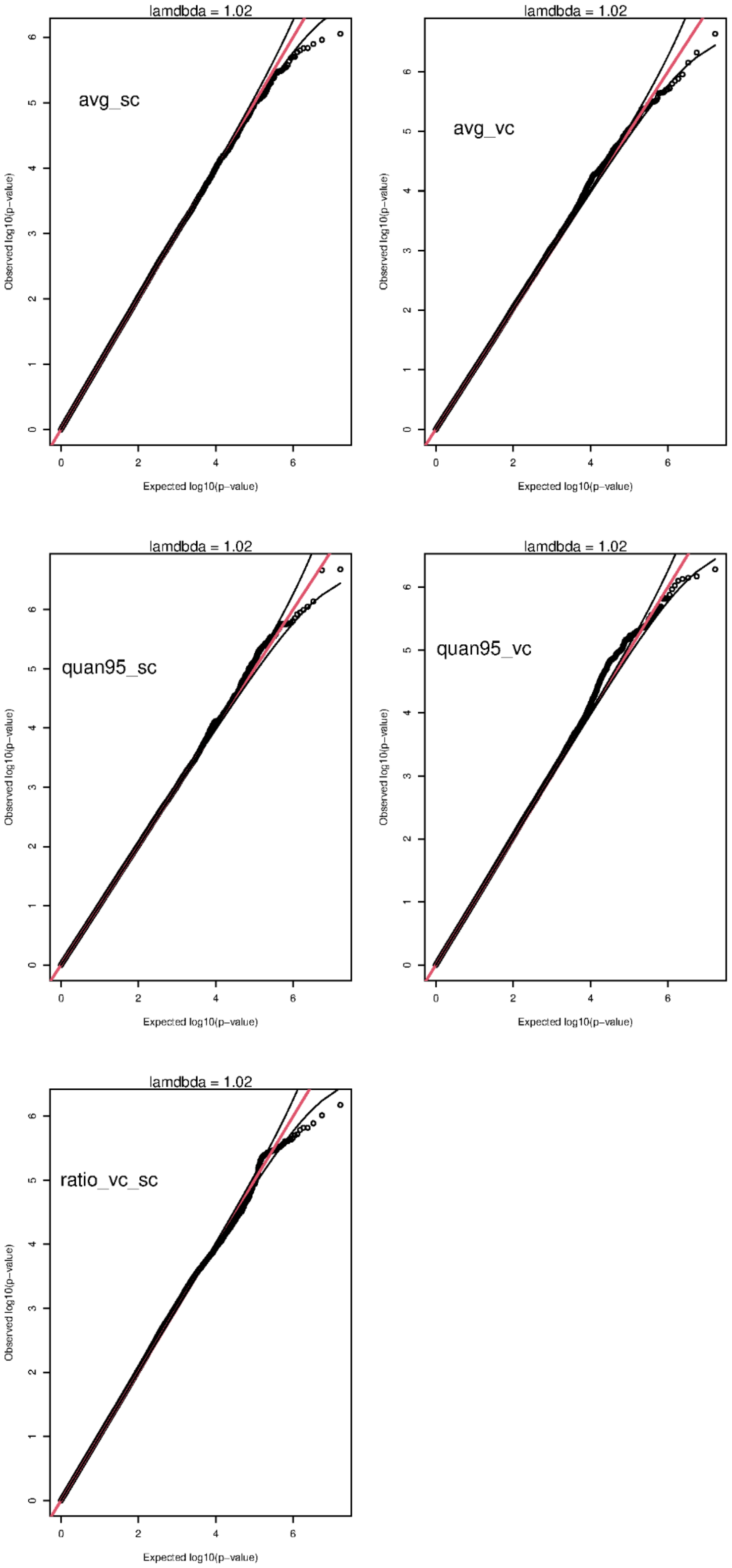


**Supplementary Figure 21.** QQ-plot for mean size of adipocytes in subcutaneous and visceral adipose tissue, 95%-quantile of adipocyte size in subcutaneous and visceral adipose tissue, and ratio of mean size of adipocytes between visceral and subcutaneous adipose tissue. Only variants present in all five cohorts are included. Males only.

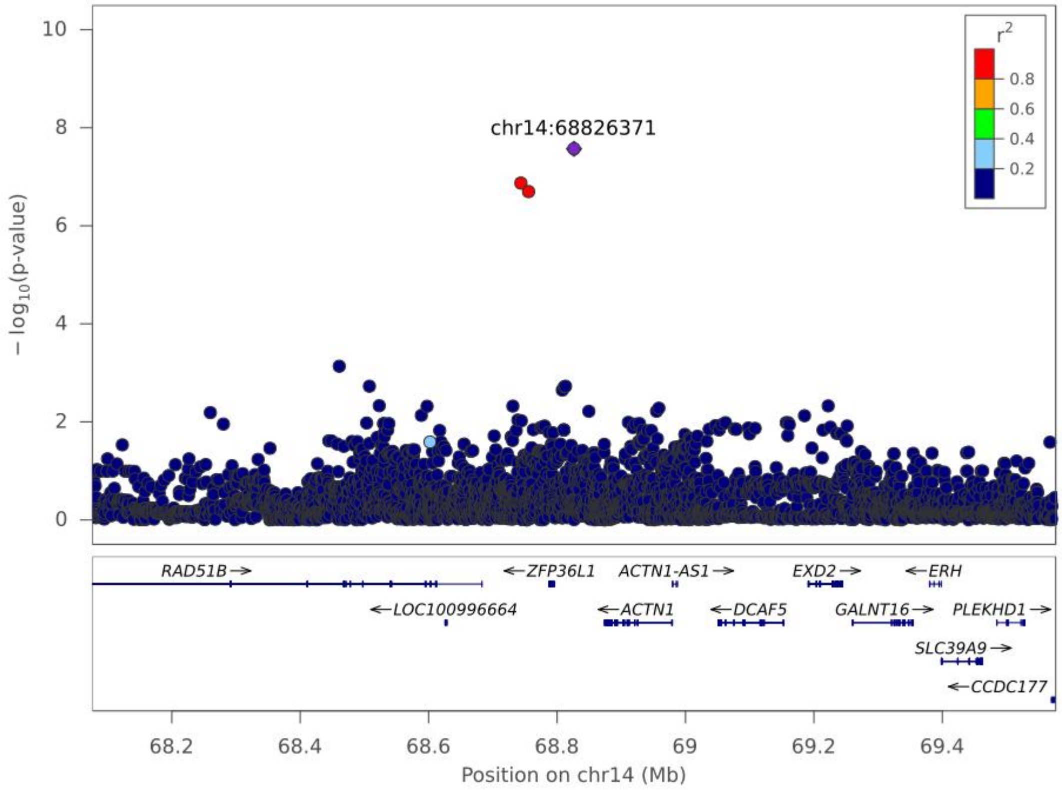


**Supplementary Figure 22.** LocusZoom plot of rs140503338 and its association with mean adipocyte size in females in subcutaneous adipose tissue. A region of +/- 750 kb has been plotted around the variant. The colour of the dots indicates the degree of LD between rs140503338 and that variant, the $R^2$ values are based on the GTEx sequence data.

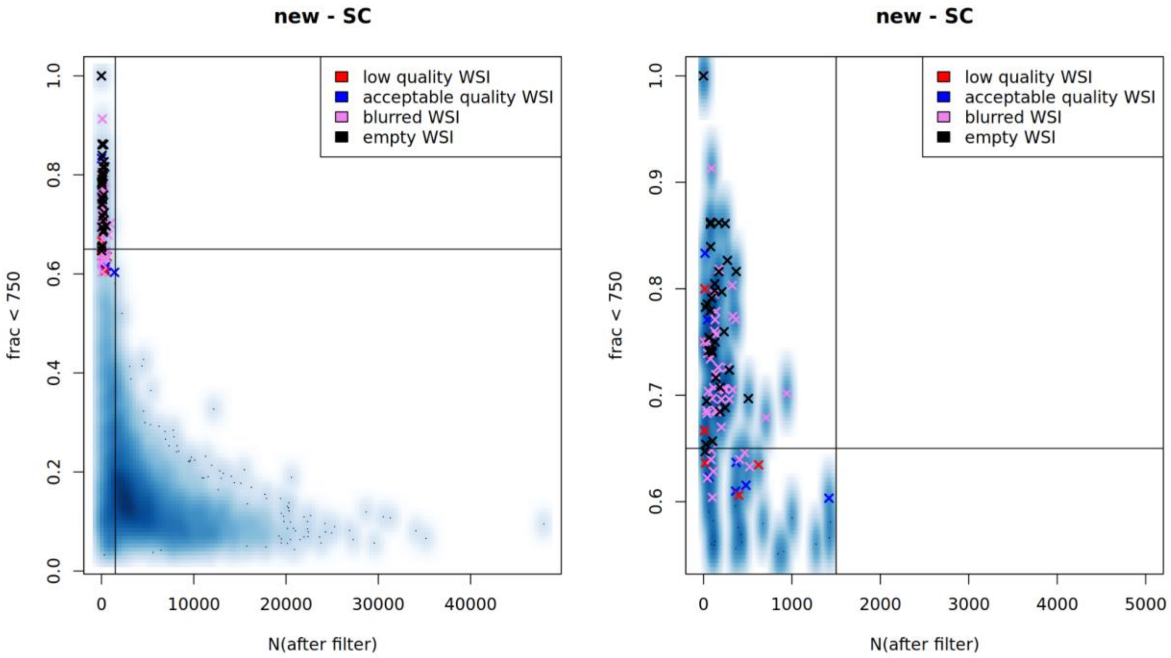


**Supplementary Figure 23.** A heatmap plot of the distribution of the fraction of adipocytes below 750 µm$^2$ (y-axis) and number of adipocytes (x-axis) for each WSI. Each cross represents a WSI that was manually inspected and categorised. The right plot is a zoomed in version of the upper left corner.

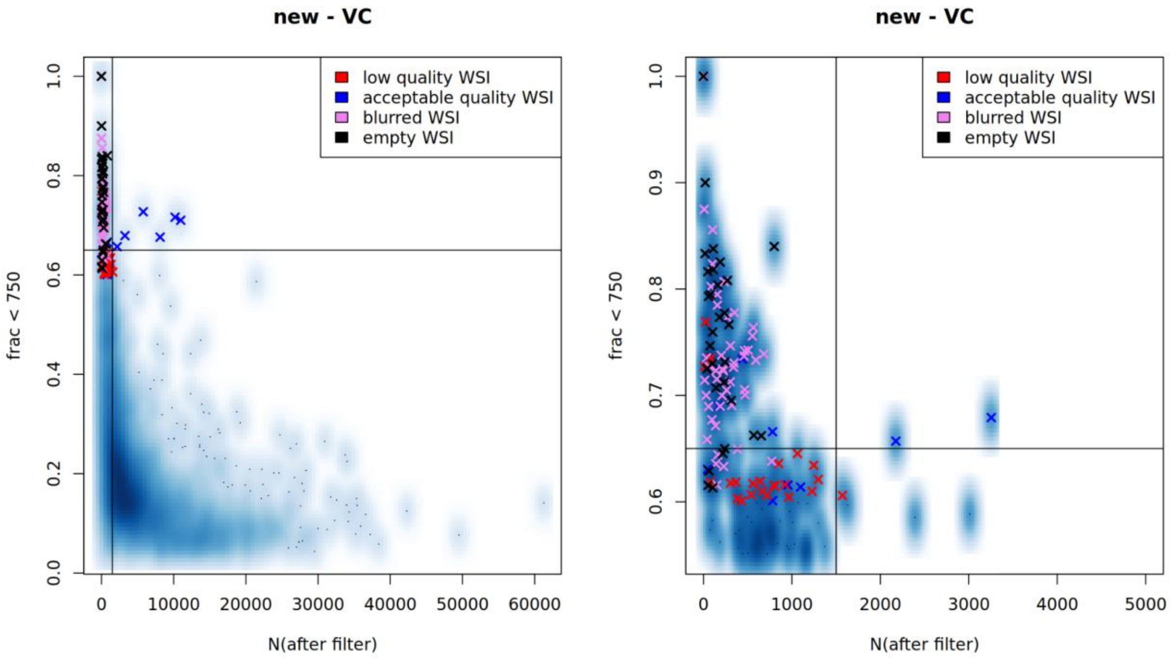

**Supplementary Figure 24.** The same as Supplementary Figure 23 however with visceral adipose tissue.

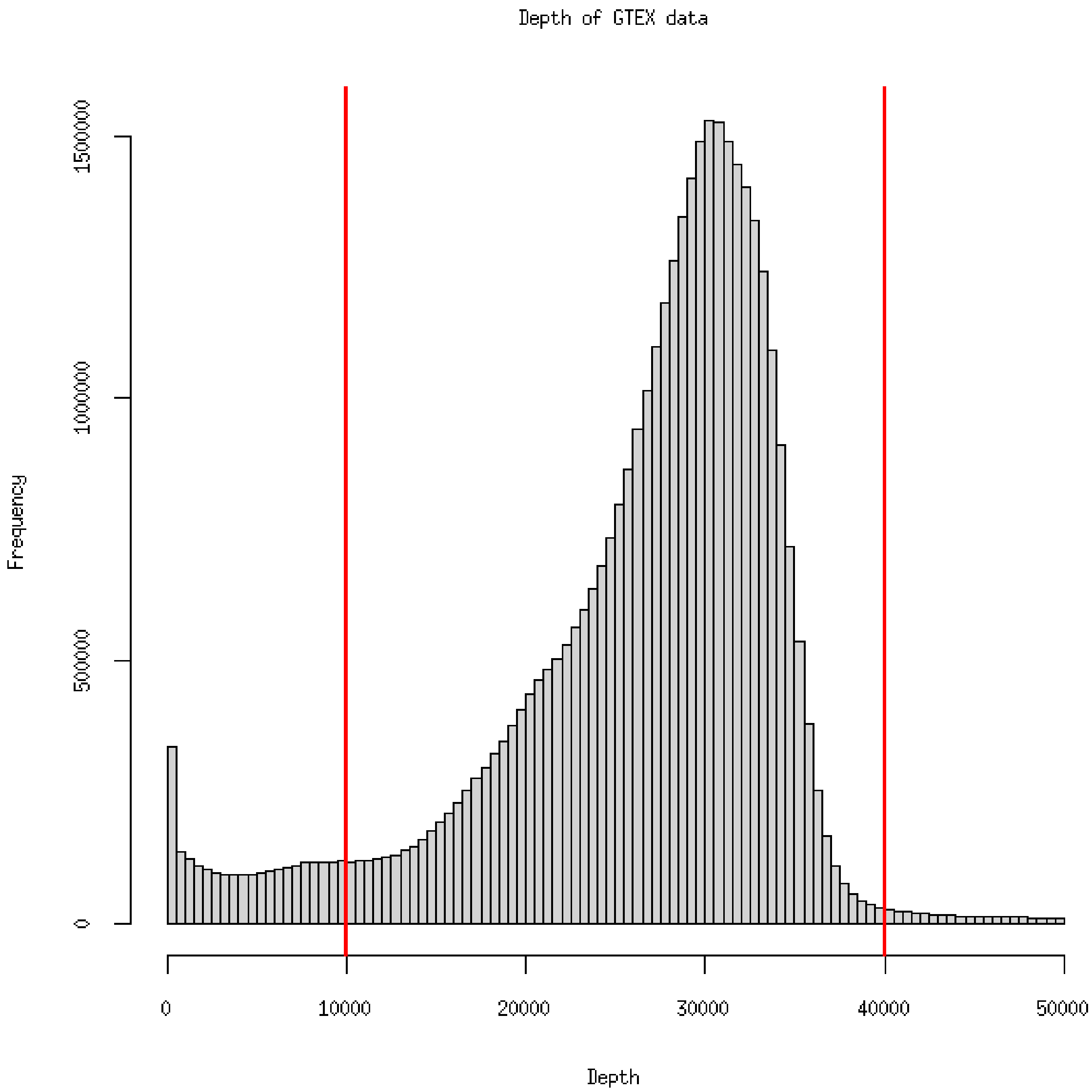


**Supplementary Figure 25.** Histogram of depth for the GTEX whole genome sequence data. The vertical red lines denote the cutoffs we have used. We only show from 0 to 50,000 depth to better visualise the distribution.

| Cohort | Individuals genotyped | Sites | Genotype Platform |
|---|---|---|---|
| Leipzig | 1,251 | 730,059 | Global Screening Array-24 v3.0 BeadChip |
| ENDOX (part I) | 56 | 655,448 | Affymetrix Axiom |
| ENDOX (part II) | 127 | 700,078 | Illumina Infinium Global Screening Array |
| Hohenheim & Munich (part I) | 175 | 730,059 | Global Screening Array-24 v3.0 BeadChip |
| Munich (part II) | 192 | 700,078 | Infinium Global Screening Array |
| GTEx | 866 | 69,763,935 | Whole genome sequenced on:<br>Illumina HiSeq 2000 machine and Illumina HiSeq X machine |

**Supplementary Table 1.** Overview of the genotyping of each cohort. GTEx donors were whole genome sequenced to a median depth of 32X. 79 GTEx donors were sequenced on an Illumina HiSeq 2000 machine, 801 donors were sequenced on an Illumina HiSeq X machine. For more information on how the GTEx cohort was sequenced see (2).

### Supplementary methods

#### Training the deep learning model

We implemented a U-net (3) based model in PyTorch (4) for the semantic segmentation of adipocytes in WSIs. The model is based on the design in (1), with added dilations for the lowest resolution layer. The model takes 1024x1024 pixel RGB images as input. A reference pixel size of 0·2500 µm was selected for this model.

The model was trained with an ADAM optimizer (5) from PyTorch, using an initial learning rate of 0·0001, decaying by a factor of 0·5 per 30 epochs, and a batch size of 2. 200 tiles were shown to the model per epoch, and the model was trained for 200 epochs. The loss function was a dice binary cross-entropy loss (half dice coefficient and half cross-entropy), and a dice coefficient metric was used to assess performance. We applied early stopping, storing the weights with the lowest validation loss.

$$Dice\ loss\ = 1 - \frac{2\sum_{n=1}^{N}\ p_n r_n + \epsilon}{\sum_{n=1}^{N}\ p_n + \sum_{n=1}^{N}\ r_n + \epsilon}$$

$$Binary\ cross\ entropy\ loss\ = -\frac{1}{N}\sum_{n=1}^{N}\ r_n log(p_n) + (1 - r_n)log(1 - p_n)$$

$$loss\ =\ 0 \cdot 5(Dice\ loss) + 0 \cdot 5(Binary\ cross\ entropy\ loss)$$

Where $p_n$ is the predicted probabilities, $r_n$ is the ground truth labels of the *n*th pixel $N$ is the total number of pixels and $\epsilon$ is a term to prevent division by 0.

During training, each input was normalised to Z-scores using the mean and standard deviation from the training data, and augmentations from the albumentations (6) Python library were applied, selecting from horizontal flip, random rotate 90 degrees, random brightness contrast, blurring and Gaussian noise. For each training tile, four random crops were sampled from the training tiles. The crops were selected from the top left, top right, bottom left and bottom right of four randomly selected tiles. The resulting crops were sampled and merged into a final tile of 1024x1024 pixels. Finally, augmentation was applied to the merged tile.

Augmentations and merging of random crops were not performed during validation. The training, validation and hold-out test split was 70%, 15% and 15%, respectively.

The dice score of our validation with the lowest loss (0·36) was 0·83 (Supplementary Figure 1), the dice score of our held-out test set was 0·87.

**Deep learning-derived phenotypes**
For prediction, we adapted the code base for HAPPY (7) implemented in Python and using PyTorch for semantic segmentation. Tiles of 1024x1024 pixels are used, starting from the top left corner of the WSI and moving left to right in rows (Figure 1). Tiles include a 256 pixel overlap with both vertically and horizontally adjacent neighbours. Tiles are inferred to be empty and excluded from further processing if all pixels are close to white or black, or if the ratio of the top decile mean pixel value to the bottom decile mean pixel value is above 0·95.

The weights for the model are from the training described in the "Training deep learning model" section.

The segmentation mask (using a 0·8 segment network confidence cutoff for saving predictions) of each tile is converted to polygons using the "measure.find_contours()" package from the scikit-image (a.k.a. skimage) Python library. The polygons are constructed using the shapely Python library.

Segmentation is performed twice on each WSI, with a pixel size of 0·2500 µm and 0·5034 µm.

After the whole WSI has been segmented, each polygon is searched for polygons in its vicinity using an STRtree (8) from the shapely library and then merged with intersecting polygons (Figure 1A).

**Post-Processing**
The merged polygon list was filtered for area greater than 316·23 $\mu m^2$ and a Polsby-Popper (PP) roundness value (9) greater than 0·6 (manually selected thresholds; prefiltered distributions and selected thresholds are shown in Supplementary Figure 2 & 3) (Figure 1A). To remove WSIs without any or very little adipose tissue, WSIs with fewer than 1500 polygons left after filtering and more than 65% of the polygons with an area below 750 $\mu m^2$ were discarded (manually selected thresholds, see Supplementary Figure 23 & 24). Additionally, we removed faulty and low-quality WSIs, upon quality control from laboratory technical staff based at Leipzig, Munich and Hohenheim.

From the filtered polygons from each individual, we calculated mean adipocyte size and upper 95%-quantile of adipocyte size, and the ratio between mean adipocyte size in visceral and subcutaneous visceral adipose tissue.

**Effect of subcutaneous adipose tissue sampling location**
In a linear model we explore if the mean adipocyte size from subcutaneous adipose tissue is different between the GTEX cohort and the other cohorts. Subcutaneous adipose tissue in GTEX was sampled from the lower leg, whereas subcutaneous adipose tissue was sampled from the stomach area in the other cohorts (1). The model was done with a categorical variable denoting if the individual is GTEX or not. The linear model was adjusted for age, sex, BMI and type 2 diabetes state (if available).

**Adipocyte size epidemiology using linear mixed model**

We used the R-package “meta” to compare mean adipocyte size, across cohorts, using a random-effects meta analysis. We chose a model with only a random intercept, comparing our effect sizes estimated in each cohort using a standard linear model.

$$\theta_i \quad = \mu + u_i$$

Where $u_i \sim N(0, \tau^2)$. The true effects are assumed to be normally distributed with mean 0 and variance $\tau^2$. If $\tau^2 = 0$, then this implies homogeneity among the true effects. Restricted maximum-likelihood is used for estimating $\tau^2$. For interpretability we also report $I^2$ estimates (in percent) how much of the total variability in the effect size estimates can be attributed to heterogeneity among the true effects ($\tau^2 = 0$ therefore implies $I^2 = 0\%$) (10).

$\theta_i$ is derived from our standard linear model.

$$y_i \quad = \theta_i + e_i$$

$\theta_i$ is the fixed effect size estimate of the *i*th factor, $y_i$ is the phenotype. We test for an association between mean adipocyte size and a whole range of epidemiological factors: depot (subcutaneous or visceral), sex, BMI, type 2 diabetes status, or age. We test in both subcutaneous and visceral adipose tissue, we additionally adjust the standard linear model with sex BMI and age, if not already included in the model. All variables were scaled to a standard score.

### Genetic Quality Control

#### Sex check

For each cohort we first updated sex from the phenotype file and we then did a sex-check (using --check-sex in plink1.9). We removed individuals with discordant genetic and phenotypic sex.

#### Variant- and individual-based QC

Looking at the distribution of missingness per variant and per individual, we elected to remove individuals with more than 2% missingness and variants with more than 5%. We removed variants with minor allele frequency below 1%. Furthermore, for Hardy Weinberg (HWE) filtering we chose a filter, based on looking at the cumulative distribution function of the chi square test statistic of the HWE test, of removing sites with $P < 1 \cdot 10^{-3}$ for the Leipzig, Munich (with Hohenheim) part I, Endox part I & II, and $1 \cdot 10^{-6}$ for Munich part II. And we removed individuals with more or less than mean +/- six units of standard deviation for heterozygosity.

For the GTEX data, where there is whole genome sequenced (WGS) data, we used the same filters as listed above. We used a threshold of $P < 1 \cdot 10^{-3}$ for removing sites, based on a HWE test. But furthermore prior to those filters, we removed sites with Variant Confidence/Quality by Depth < 5, and sites with total depth < 10,000 or > 40,000 (see Supplementary Figure 25). Furthermore, we only kept sites that passed the initial QC of the WGS data (FILTER is equal to “PASS”). And removed individuals with chromosomal abnormalities, based on information provided by GTEX.

#### Relatedness

The relatedness analysis was run using --genome in plink 1.9 (11) with --ppc-gap 0. First the data was LD pruning using --indep-pairwise 100 10 0.2. We removed one individual from pairs of individuals with PI_HAT (P(IBD=2) + 0·5*P(IBD=1), IBD=identical by descent) above 0·9, as we deemed these were duplicate individuals.

#### PCA

First the script HRC-1000G-check-bim.pl was run, making sure the SNP-chip variants all are on the plus strand. Also it removes variants that differ from their reference in one of the following ways, do not have the same alleles, where allele frequency difference is greater than 0·2, or variants that are not found. Each SNP-chip dataset was merged with 1000 genomes data (12), removing related individuals from 1000 genomes, and outliers. After merging a filter of removing variants with missingness > 1% and MAF < 5 % was applied, and LD pruning (same options as above). For each cohort, non-Europeans were removed from the cohort. The non-Europeans were identified by drawing a box around the Europeans from the PCA plot of each SNP-chip merged with the 1000 genomes data. This left this many individuals in each cohort. Leipzig - N=1201 (removing 13 based on the PCA), Munich - N=183 (removing 7 based on the PCA), Munich & Hohenheim - N=149 (removing 25 based on the PCA), Endox I (affy) - N=56 (removing 0 based on the PCA), Endox II (Ilm) - N=127 (removing 0 based on the PCA).

8. Leutenegger ST, Institute for Computer Applications in Science and Engineering. STR: A Simple and Efficient Algorithm for R-Tree Packing. Institute for Computer Applications in Science and Engineering, NASA Langley Research Center; 1997. 34 p.

9. Polsby D, Popper R. The Third Criterion: Compactness as a Procedural Safeguard Against Partisan Gerrymandering. 2015 Oct 16 [cited 2024 Mar 18]; Available from: https://digitalcommons.law.yale.edu/ylpr/vol9/iss2/6

10. Viechtbauer W. Conducting Meta-Analyses in R with the metafor Package. J Stat Softw. 2010 Aug 5;36:1–48.

11. Chang CC, Chow CC, Tellier LC, Vattikuti S, Purcell SM, Lee JJ. Second-generation PLINK: rising to the challenge of larger and richer datasets. Gigascience. 2015 Feb 25;4:7.

12. 1000 Genomes Project Consortium, Auton A, Brooks LD, Durbin RM, Garrison EP, Kang HM, et al. A global reference for human genetic variation. Nature. 2015 Oct 1;526(7571):68–74.
